# Goal-Directed Control in Schizophrenia: Loss-Biased Engagement of the Anterior Cingulate Relates to Negative-Symptom Outcomes

**DOI:** 10.1101/2025.07.02.25330758

**Authors:** Wolfgang Omlor, Giacomo Cecere, Akhil Ratan Misra, Gao-Yang Huang, Xiaoyan Wu, Victoria Edkins, Giulio Pergola, Boris Quednow, Philippe Tobler, Philipp Homan

**Author notes:** Correspondence concerning this article should be addressed to: Wolfgang Omlor, MD, PhD, Psychiatric Hospital, University of Zurich, Lenggstrasse 31, 8032, Zurich, Switzerland., Philipp Homan, MD, PhD, Psychiatric Hospital, University of Zurich, Lenggstrasse 31, 8032 Zurich, Switzerland. These authors contributed equally (Wolfgang Omlor, Giacomo Cecere, Akhil Ratan Misra).

## Abstract

**Background and Hypothesis:** Individuals with schizophrenia spectrum disorders (SSD) often show diminished reward pursuit, whereas loss avoidance is relatively preserved. The neural mechanisms of this dissociation and its relation to negative symptoms remain unclear. We hypothesized that in SSD, cognitive resources are preferentially directed toward avoiding losses rather than pursuing rewards, potentially limiting reward processing and contributing to negative symptoms.

**Study Design:** Using computational modeling of behavior during a two-stage decision task which distinguished between goal-directed (model-based) and habitual (model-free) strategies under reward and loss conditions, we studied 42 stable individuals with SSD and 48 healthy controls (HC) during functional magnetic resonance imaging.

**Study Results:** In individuals with SSD, model-based control was shifted toward loss avoidance relative to HC, with corresponding changes in prefrontal circuitry. In anterior cingulate, orbitofrontal, and dorsolateral prefrontal regions, individuals with SSD showed increased activation during model-based control in the loss condition. Within this group, loss-biased activation in the right anterior cingulate region was associated with anhedonia. In 25 patients with available follow-up data, loss-biased activation in the right anterior cingulate region at baseline was prospectively related to worsening of motivation and social engagement over the subsequent year.

**Conclusions:** Our findings suggest that, compared to HC, those with SSD allocate their limited cognitive resources more toward loss avoidance relative to reward pursuit. The association between loss-biased anterior cingulate engagement and anhedonia supports a neurocomputational account of diminished pleasure in psychotic disorders, with potential implications for developing motivation-targeted treatments and early prediction of negative-symptom worsening.

## Introduction

How does the mind decide when it is worth investing cognitive effort to chase a reward versus avoid a loss? Everyday choices—double-checking a tax return to avoid a penalty or staying late at work to secure a bonus—force the brain to weigh potential gains against possible costs^1^. In schizophrenia spectrum disorders (SSD), which are marked by profound disturbances in cognition and goal-directed behavior^2–4^, this balance between reward pursuit and loss avoidance appears to be altered and related to negative symptoms^5^. Strikingly, the drive to pursue rewards is blunted, whereas the capacity to avoid losses remains intact or is even heightened^5, 6^. This asymmetry raises the possibility that cognitive resources for goal-directed behavior are preferentially used to avoid losses at the expense of reward pursuit^7, 8^. Yet this cognitive resource-reallocation hypothesis has not been tested within a computational framework that separates goal-directed from habitual control and manipulates motivational context in individuals with SSD.

The two-stage reinforcement-learning task, coupled with computational modeling, allows researchers to quantify the balance between two principal decision processes that place different demands on cognitive resources: model-based control is goal-directed, forward-looking, cognitively demanding, slow, and flexible, whereas model-free control is habitual, backward-looking, resource-efficient, fast and rigid^9–30^. The two-stage decision task has been used to demonstrate that compulsive conditions, including obsessive–compulsive disorder and substance dependence, are associated with reduced model-based control in reward contexts, but not in loss contexts^31, 32^. Because model-based control hinges on cognitive resources^14, 15^ and dopaminergic signaling^33–36^, both often compromised in psychotic disorders^3^, individuals on the schizophrenia spectrum are expected to exhibit impaired model-based learning^4^. Indeed, in reward variants of the two-stage decision task, individuals with SSD have shown impairments in model-based control^37^.

It remains unclear, however, how reward and loss contexts modulate the balance between model-based and model-free control in individuals with SSD. Resolving this knowledge gap is essential to determine whether psychosis shifts goal-directed cognitive control toward loss avoidance at the expense of reward pursuit, and whether this distortion drives reward devaluation and negative symptoms. We therefore aimed to test whether model-based control and its neural implementation are redirected from reward pursuit to loss avoidance in individuals with SSD, and how this reallocation relates to distinct symptom domains. Guided by evidence that SSD entail restricted cognitive capacity^2, 3, 38^, value representation is dependent on cognition^7, 8, 39^, and reward pursuit is attenuated relative to loss avoidance^5^, we derived a set of specific questions for behavior, neural engagement and their clinical correlates. We hypothesized that limited cognitive resources in psychotic disorders are preferentially allocated to goal-directed avoidance of negative outcomes rather than reward pursuit, and therefore asked whether:

(i) individuals with SSD would show relatively preserved model-based control when choices avert losses alongside attenuated model-based behavior when they secure gains; (ii) any such goal-directed control asymmetry would be accompanied by a corresponding neural signature—specifically, stronger loss-than reward-related activation in three prefrontal hubs implicated in model-based computations^4^: the anterior cingulate cortex (ACC) for allocating cognitive resources according to the expected value of control^40–43^, the orbitofrontal cortex (OFC) for prospective value representation^5, 44, 45^, and the dorsolateral prefrontal cortex (DLPFC) for planning and working memory^46, 47^; and (iii) the magnitude of loss-biased recruitment of model-based control regions would be associated with negative-symptom severity. Given the link between blunted reward learning with preserved loss avoidance and negative symptoms in SSD^5^, we considered that this motivational asymmetry could reflect aberrant allocation of goal-directed control that is linked to negative symptoms.

To address these questions, we combined functional magnetic resonance imaging (fMRI) with the canonical two-step sequential decision task^9^ (**Fig. 1a**) in medicated, clinically stable individuals with SSD and healthy controls (HC). Separate reward and loss versions of the task were used during fMRI to map prefrontal activation in the ACC, DLPFC, and OFC (**Fig. 1b**) during model-based control. This design allowed us to quantify context-dependent shifts between model-based and model-free control and to characterize their neural signatures within these regions (**Fig. 1c**). In a subset of participants with SSD who had longitudinal clinical data, we investigated in an additional explorative analysis whether context biased activation during model-based control predicted one-year follow-up levels of negative-symptom–related domains. This mechanistic–prognostic approach links circuit-level computations to both cross-sectional symptom variation and the longitudinal course of negative-symptom–related outcomes.

**Figure 1:**
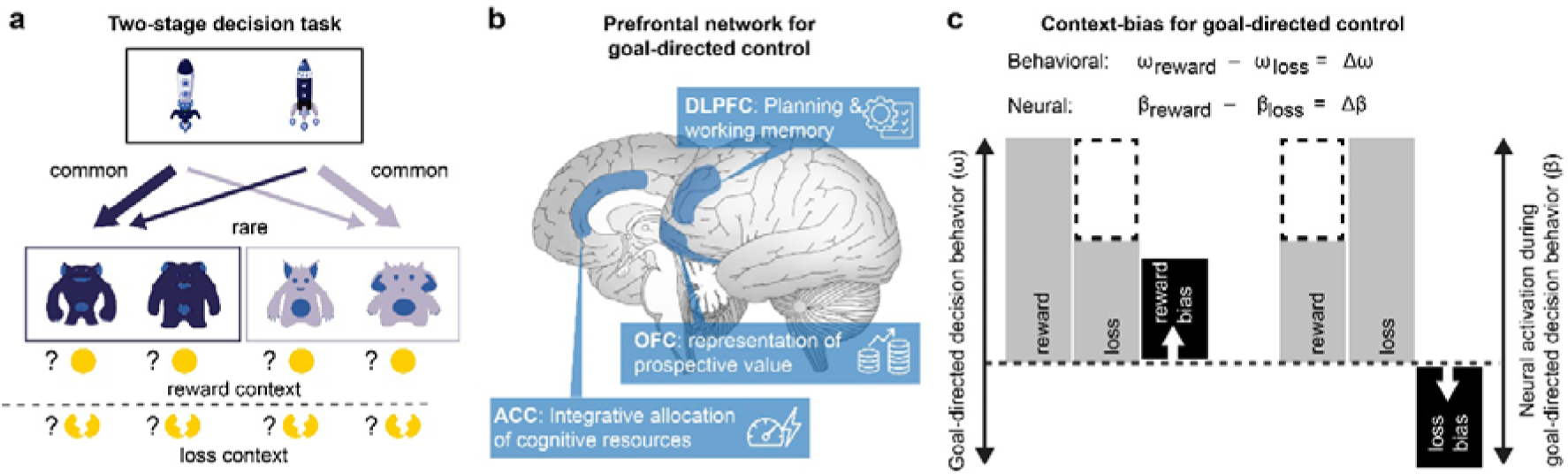
Experimental setting and modeling hypothesis. **(a)** *Two-stage decision-making task.* Adapted from Decker, Otto, Daw, Hartley ^49^. Participants made two consecutive choices to either earn play coins (*reward context*) or avoid losing play coins (*loss context*). Probabilistic transitions between the first and second stages (common vs. rare) allowed for the quantification of model-based and model-free control. In stage 1, each spaceship was associated predominantly (70% probability,’common’ transitions) with one of the two alien pairs in stage 2, and less frequently (30% probability,’rare’ transitions) with the other pair. **(b)** Schematic of the human prefrontal network supporting model-based reinforcement learning. Dorsolateral prefrontal cortex (DLPFC) supports planning and working-memory processes; orbitofrontal cortex (OFC) encodes prospective option values; anterior cingulate cortex (ACC) integrates these signals to allocate cognitive control. **(c)** Schematic showing the computation of context biases in goal-directed control. For each participant, the model-based weight ω and the task-evoked BOLD parameter β were estimated separately in *reward* and *loss* contexts (grey bars). Context bias was defined as the difference Δω = ω_reward_– ω_loss_ for behavior and Δβ = β_reward_– β_loss_ for neural activation. Positive Δ indicates a *reward bias* (black bar with arrow pointing up), whereas negative Δ indicates a *loss bias* (black bar with arrow pointing down).

**Figure 2:**
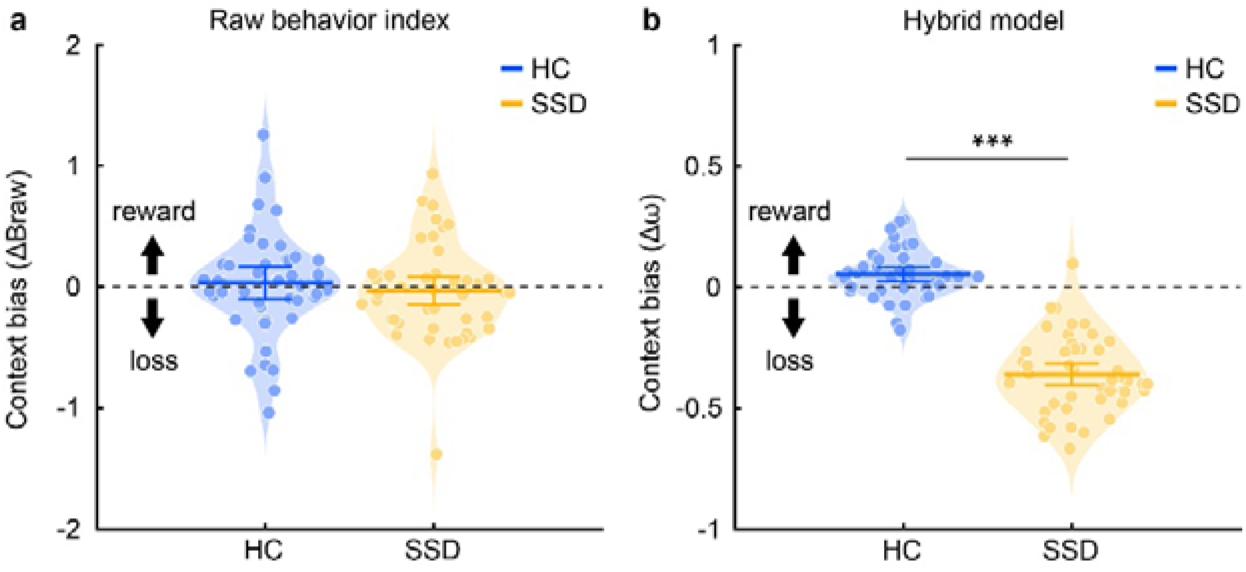
Analysis of decision behavior: **(a)** *Simplified raw behavior scores:* Regarding the relative contribution of model-based learning, healthy controls (HC blue) exhibited a numerically higher mean contribution of model-based control in the reward relative to the loss context, whereas individuals with schizophrenia spectrum disorders (SSD) showed a numerically higher mean contribution of model-based control in the loss relative to the reward context. When the context bias of raw behavior scores was compared between groups the difference was not significant. **(b)** _ω_*-scores from the modeling analysis:* Regarding the relative contribution of model-based learning, the ω-scores indicated a significant reward bias in HC and a significant loss bias in individuals with SSD. In addition, the context bias of ω-values differed significantly between groups. Mean and 95% confidence intervals are shown. Asterisks indicate *p* < 0.05.

## Methods

### Participants

50 individuals with SSD were recruited from the University Hospital of Psychiatry in Zurich, and 48 HC were recruited through local advertisements. Diagnoses were established with a structured clinical interview and according to ICD-10 criteria. Eligible patients met criteria for SSD and were within five years of their first psychotic episode. In addition, eligible patients showed no evidence of substance-induced psychosis. All patients were clinically stable, in remission, and receiving antipsychotic medication at the time of testing. HC had no lifetime history of psychiatric or neurological disorders. The study was approved by the Cantonal Ethics Committee of Zurich, and all procedures adhered to its guidelines and complied with the Declaration of Helsinki. All participants provided written informed consent prior to participation, after receiving a complete description of the study. The final analysis sample comprised 48 HC and 42 participants with SSD. Eight participants with SSD were excluded a priori for quality reasons: four for insufficient task engagement and four for excessive head motion or incomplete fMRI acquisition (details below).

### Clinical Assessments

Symptoms were assessed using several rating scales. The Brief Negative Symptom Scale (BNSS) was used to assess the different negative symptom domains of the apathy (avolition, asociality, anhedonia) and diminished expression (blunted affect, alogia) dimensions^48^. To complement this domain-specific measure, overall psychopathology was evaluated with the Brief Psychiatric Rating Scale (BPRS). Detailed cognitive functioning was characterized with the MATRICS Consensus Cognitive Battery (MCCB). All participants were required to pass a urine drug screen and a breathalyzer test. In addition, AMDP clinical ratings (0–3 scale; 0 = absent, 1 = mild, 2 = moderate, 3 = severe) were obtained at baseline and at follow-up assessments at 3, 6, 9, and 12 months if individuals with SSD were available. Although AMDP ratings provide a standardized, time-efficient clinician-rated assessment across a broad range of psychopathological symptoms, ratings were available only for a subset of patients (n=25) who attended outpatient follow-up visits or were clinically stable during subsequent hospitalizations, and within this subset they were not consistently obtained at all prespecified time points (**Supplementary Tables 1–4**). As apathy-related proxies (avolition, asociality, anhedonia), we used the AMDP items amotivation and social withdrawal (AMDP does not include a direct anhedonia item). As diminished-expression proxies (blunted affect, alogia), we used constricted affect and affective rigidity (AMDP does not include a direct alogia item).

### Task Design

We used the two-stage task as described in previous research^9^. In total, participants completed 400 trials (200 trials per context) with 3 seconds allocated for each choice. The interval between first and second-stage stimuli was 1 second, followed by 1 second of feedback presentation after the second-stage choice. The sequence of events in each trial is illustrated in **Fig. 1a**. We employed the simplified version of the two-stage decision task described by Decker et al.^49^, which replaces abstract fractal cues with easily discriminable “spaceship” and “alien” stimuli. This adaptation with an embedded story line lowers perceptual and working-memory demands, accommodating the cognitive impairments often observed in SSD, and provides a more intuitive, engaging environment for participants.

Each trial included two stages: In the first stage, participants used an MRI-compatible button box to select one of two spaceships. Choices had to be made within 2 seconds; otherwise, the trial was aborted. Once a choice was made, the selected option moved to the top of the screen, while the unselected option disappeared. In the second stage, participants were presented with one of two new states (two red or two purple aliens) each containing two additional options. The participants selected one option, and the outcome was displayed. In the reward setting, the outcome was either a monetary reward (represented by a coin) or no reward (represented by a red cross). In the loss setting, the outcome was either a monetary loss (represented by a broken coin) or no loss (represented by a green cross). Trials were separated by an inter-trial interval of randomized duration, averaging approximately one repetition time (TR). The second-stage state presented was probabilistically determined by the first-stage choice, based on the transition scheme shown in **Fig. 1a**. The position (left or right) of the options at each state was randomly varied between trials. Reward probabilities for each bottom-stage option were dynamic, fluctuating with independent Gaussian noise^9^. This dynamic structure encouraged ongoing learning. The order of reward and lossLtrial blocks (a block of 200 trials in each context) was randomized for each participant.

Before the fMRI task, participants completed a computerized training session to understand the task structure. In this training session, participants completed 50 practice trials. They were informed that reward probabilities would vary throughout the task, but the transition probabilities between the first and second stages would remain fixed. Participants were also told that each first-stage choice was predominantly associated with one of the two second-stage states, and the specific mapping was disclosed. Participants were informed that they would receive 50 CHF for task completion. Four participants with SSD did not engage with the task (always selected the same response option/pressed the same button during one stage)—and were therefore excluded from further analysis.

### Behavior analysis

To examine learning effects without presupposing a particular reinforcement-learning model, we first asked how events on the previous trial shaped choice on the subsequent trial. We computed a stay-probability (StP) index based on the combinations of transition type (common/rare) and outcome (reward/loss). From this, we derived straightforward and intuitive “raw” metrics of model-based (MB_raw_) and model-free (MF_raw_) control (**Supplementary Fig. 1**). Model-based and model-free raw scores were calculated separately for reward and loss contexts as follows:

### Reward context

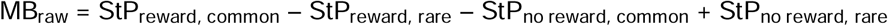

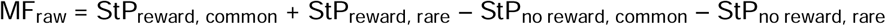

*Loss context:*

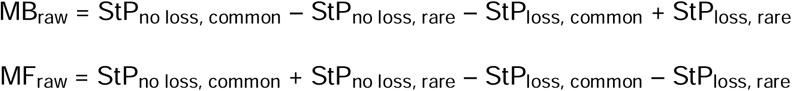

Separately for each context, we defined the balance between model-based and model-free control (B_raw_) as the difference between the respective raw scores, providing a straightforward and intuitive metric to quantify their relative contributions:

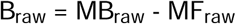

To quantify the context-dependent difference in the deployment of model-based versus model-free control, we defined the raw context bias (CB_raw_) as the difference between the B_raw_ scores obtained in the reward and loss contexts, computed as follows:

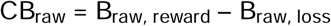

Computational modeling provides a powerful framework for systematically quantifying latent cognitive processes underlying learning tasks, allowing the precise characterization of mechanisms that cannot be directly inferred from behavioral data alone^9, 50, 51^. Therefore, we additionally deployed a hierarchical Bayesian reinforcement-learning model that incorporates the full choice history and individual learning rates, providing a generative, mechanistically explicit estimate of model-based versus model-free control. We fitted a hybrid reinforcement-learning model that integrates two algorithms: (i) model-based control, which computes action values through prospective (recursive) planning over the task’s transition structure; and (ii) model-free control, which updates action values via reward-prediction errors, strengthening actions that are rewarded and weakening those followed by loss^9, 52^. A key parameter, denoted as ω, determines the balance between these systems in shaping first-stage choices, where their predicted values diverge most. This weighting parameter is fixed across trials, with ω = 1 reflecting fully model-based control, and ω = 0 indicating purely model-free influence. Further model specifications and fitting procedures are detailed in the supplementary materials. Similar to ω, B_raw_ quantifies the relative contribution of model-based versus model-free learning, with higher values indicating greater reliance on model-based control. To assess the correspondence between the two metrics, we computed their Spearman correlation. To quantify the context-dependent difference in the deployment of model-based relative to model-free control, we defined the context bias (CB) as the difference between the ω scores obtained in the reward and loss contexts (**Fig. 1c**):

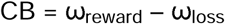

Hierarchical Bayesian parameter estimation was implemented through Stan (v. 2.18.2). Prior research has demonstrated that hierarchical parameter estimation provides more stable and reliable estimates compared to individual-level estimation^51^. Posterior distributions were approximated using the Markov Chain Monte Carlo (MCMC) algorithm. Each model was run with four chains, comprising 1000 burn-in samples and 5000 post-burn-in samples. Mixing and convergence of the chains to stationary values were assessed by computing the convergence index (L) and through visual inspection^51^.

For the two-stage decision task, we utilized the *ts_par7* model that is implemented in *hBayesDM package in R* and incorporated both reward and loss contexts^51^. This model quantifies trial-by-trial decision-making dynamics, using seven parameters: α_1_ (learning rate in stage 1), β_1_ (inverse temperature in stage 1), α_2_ (learning rate in stage 2), β_2_ (inverse temperature in stage 2), pi (perseverance), ω (balance between model-based and model-free learning) and lambda (eligibility trace). To determine the optimal specification, we also fitted four-and six-parameter variants of the two-stage reinforcement-learning model and evaluated their performance using the leave-one-out information criterion (LOOIC)^51^. LOOIC favored the seven-parameter model over both the four-and six-parameter models (**Supplementary Table 5**). Following the approach of previous work^53^, we assessed the identifiability of the seven-parameter two-stage reinforcement-learning model by simulating datasets from 100 synthetic agents, separately for the reward and loss settings. Each agent completed 200 trials (matching the empirical reward and loss blocks), and the same model was re-fit at the individual (non-hierarchical) level. Recovery was quantified as the Spearman rank correlation between true and recovered parameters across agents (**Supplementary Figs 2 and 3**).

**Figure 3:**
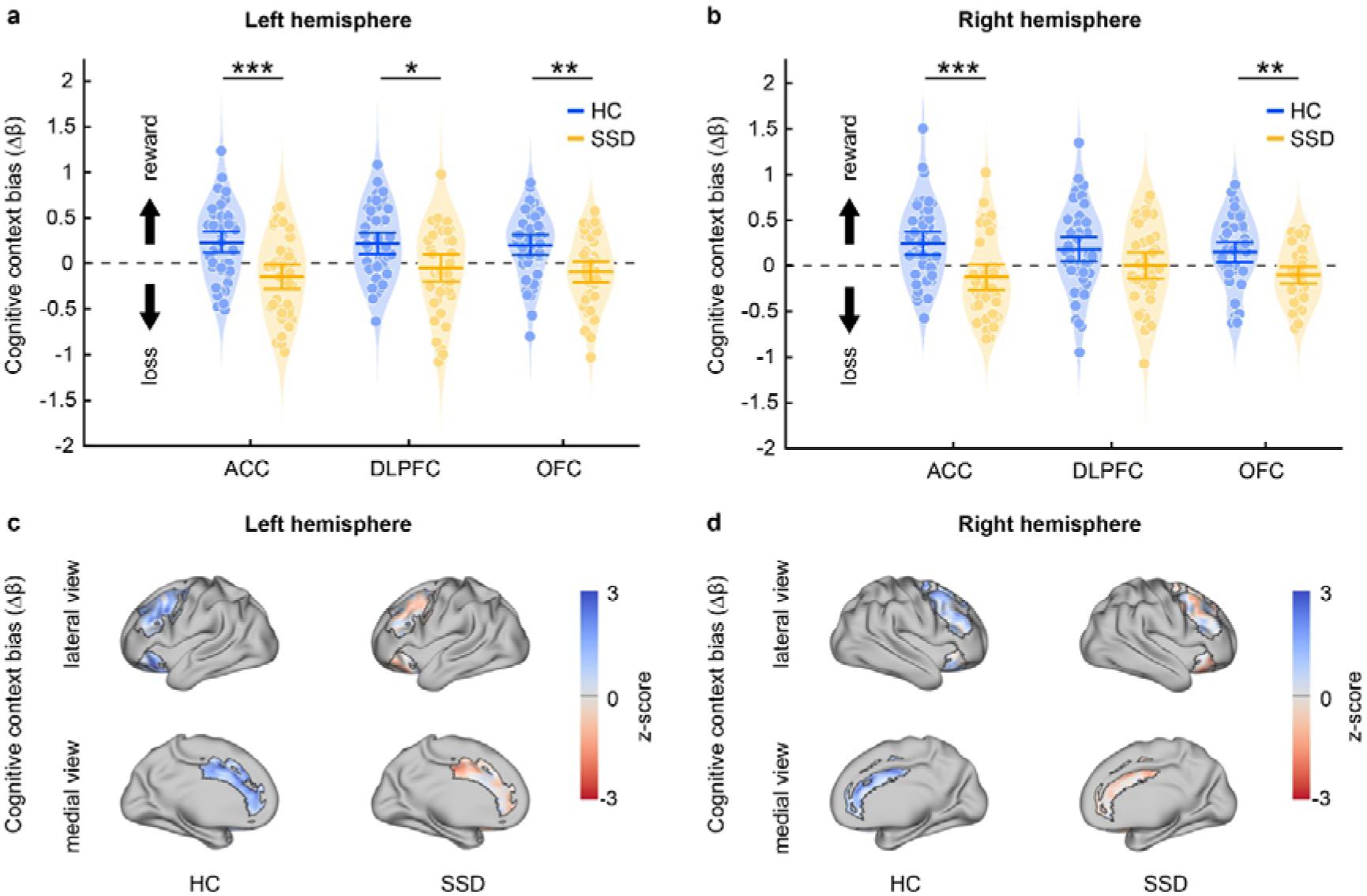
Context-dependent BOLD activation during model-based control: (**a-b**). The context bias of activation in the anterior cingulate cortex (ACC), dorsolateral prefrontal cortex (DLPFC) and orbitofrontal cortex (OFC) for healthy controls (HC, blue) and individuals with schizophrenia spectrum disorders (SSD, orange) is shown, plotted separately for the left and right hemispheres. Symbols indicate group means and 95 % confidence intervals. Asterisks indicate p < 0.05. (**c-d**) Brain maps displaying context-biased activation within ACC, DLPFC, and OFC masks, separately for the left (c) and right hemisphere (d). Maps are presented unthresholded for visualization purposes.

### MRI processing

Imaging was acquired on a Philips Achieva 3LT scanner with a 32-channel head coil during resting state and during the reward and loss versions of the two-stage decision task. Preprocessing followed an fMRIPrep-based workflow. Full acquisition parameters, preprocessing steps, quality-control criteria, and exclusions are detailed in the Supplementary Material. Four SSD participants were excluded from fMRI analyses: two for excessive head motion and two for scanner malfunction resulting in incomplete data.

### Analysis of fMRI Data

To investigate neural activity associated with task conditions, an event-related analysis was implemented using generalized linear modeling in FSL-FEAT. The analysis was based on the timeseries of Reward Prediction Errors (RPEs) as generated from the simulation of the model over each subject’s task choices. This approach followed a framework inspired by Daw, Gershman, Seymour, Dayan, Dolan ^9^, concentrating on prediction errors occurring at two critical moments within each trial: the onset of second-stage stimuli and the onset of reward delivery.

For the first-level analysis, two onset regressors were included in the model (i) first-stage onset (ii) second stage and feedback onset time incorporated into the model as a single regressor. The first onset regressor was parametrically modulated by the value of the chosen first-stage action and its partial derivative with respect to the ω-parameter from computational modeling (see also Supplementary Information). On the other hand, the second onset regressor was modulated by model-free prediction error and by the difference of model-free and model-based reward prediction error and its partial derivative with respect to the ω-parameter. Note that this difference regressor is zero at reward delivery as both algorithms converge at this point. To avoid confounding of the neural results due to activity differences between these two time points per se, within subject mean centering of the difference regressor was carried out. The analysis also included nuisance onset at feedback. Additionally, to correct for artefacts caused by physiological noise we included 6 motion parameters as estimated by fMRIPrep: a regressor for framewise displacement and an additional five noise regressors calculated using anatomical CompCor. To quantify context-dependent differences in neural activation associated with model-based versus model-free control, we defined the neural context bias (nCB) as the difference between β coefficients for model-based reward prediction errors estimated in reward versus loss contexts (**Fig. 1c**):

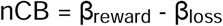

We defined activation as’loss-biased’ in a given group when the significant group difference in nCB indicated a numerical reward bias in the comparison group shifting toward a numerical loss bias in the target group. Conversely, we defined activation as’reward-biased’ when a numerical loss bias in the comparison group shifted toward a numerical reward bias in the target group.

### Region of interest and whole brain analysis

Guided by convergent evidence across mammalian studies, we selected the three prefrontal regions most consistently implicated in model-based control—the OFC, ACC and DLPFC— as primary regions of interest (ROIs) to examine their context-specific activation patterns^4, 40, 43, 44, 46^. Although some studies implicate additional regions such as the hippocampus in model-based control^54^, a quantitative whole-brain meta-analysis in human participants found no consistent hippocampal involvement in standard two-stage decision tasks and concluded that the hippocampus plays a substantial role primarily when tasks include explicit spatial relations between decision stages^44^. We excluded striatal regions as primary ROIs because they encode a mixture of model-based and model-free signals^9, 44^, whereas our focus was on regions selective for model-based control. Our results also confirm and corroborate earlier evidence^9^ that the ventral striatum encodes a mixture of model-based and model-free value signals (**Supplementary Fig. 4**). ROI masks were taken from the Harvard-Oxford cortical atlas to ensure anatomical precision (see also Supplemental Data). Due to the absence of explicit labels for certain prefrontal subregions in the Harvard–Oxford atlas, the middle frontal gyrus was used as a proxy for the DLPFC^55^. Each ROI was further divided into left and right hemispheres to allow for lateralized analysis. These resulting binary masks were applied to extract the mean BOLD activity within each region for subsequent analyses. This anatomically guided approach ensured consistency across subjects and enhanced interpretability of ROI-based neural activation patterns during task performance. We additionally examined the spatial distribution of activation within each ROI. This analysis provided a finer-grained view of how each region supports model-based control in the reward relative to loss contexts. Because our ROI analysis was hypothesis-driven and therefore might overlook regions that contribute to model-based control, we also conducted an exploratory whole-brain analysis.

### Statistical analyses

Potential differences between individuals with SSD and healthy participants were assessed using two-tailed Wilcoxon rank-sum tests. Within-group differences across contexts (e.g., omega parameters in reward versus loss contexts within the schizophrenia group) were evaluated using two-tailed Wilcoxon signed-rank tests. Associations between context-dependent ROI activation bias/behavior bias and different variables (negative symptom domains, age, sex, chlorpromazine equivalents, illness duration, occupancy of 5-HT_2A_ and D_2_-receptors^56^ were initially explored with Spearman’s rank correlations to obtain robust estimates. Significant associations were further examined using general linear models (GLMs), adjusting for age, sex, antipsychotic dose, and illness duration. ROIs were further examined in a longitudinal analysis in which we tested whether baseline context-biased activation predicted the follow-up course of negative-symptom–related features derived from AMDP ratings. To aid interpretation, we recoded the AMDP items so that higher scores indicate better functioning and related them to context-biased activation (positive = reward bias, negative = loss bias): motivation (inverse of reduced drive), social engagement (inverse of social withdrawal), affective range (inverse of constricted affect), and affective flexibility (inverse of affective rigidity). In the longitudinal AMDP analysis (see above), we computed per domain (e.g. motivation) the mean follow-up severity (FU; arithmetic mean across all available assessments between 3 and 12 months) for each participant with at least 1 follow-up. We also recorded K, the per-participant number of available follow-up assessments (range 1–4), to account for unequal follow-up density. To investigate the prospective association of context-biased ROI activation with follow-up features that are related to negative symptoms, an ANCOVA was used in the SSD group:

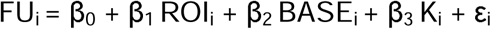

where FU_i_ is the mean 3-12 months domain severity for participant i, ROI_i_ is the context bias of participant i in the given ROI at baseline, BASE_i_ and K_i_ are mean-centered baseline level and follow-up count, and ε_i_ ∼ N(0, σ^2^). We report unstandardized coefficients (β) with 95% confidence intervals, model R^2^, and partial R^2^ for the ROI term (unique variance beyond baseline and K). Added-variable plots (**Fig. 4c-f**) display follow-up scores adjusted for baseline and K versus rACC context bias. Two-sided tests were used. The association between true and recovered model parameters was investigated with Spearman’s rank correlations. For all analyses involving multiple comparisons, p-values were corrected using FDR correction according to Benjamini-Hochberg. Statistical significance was defined as a FDR-corrected *P*L<L0.05. Unless noted otherwise, reported p-values are FDR-adjusted.

**Figure 4:**
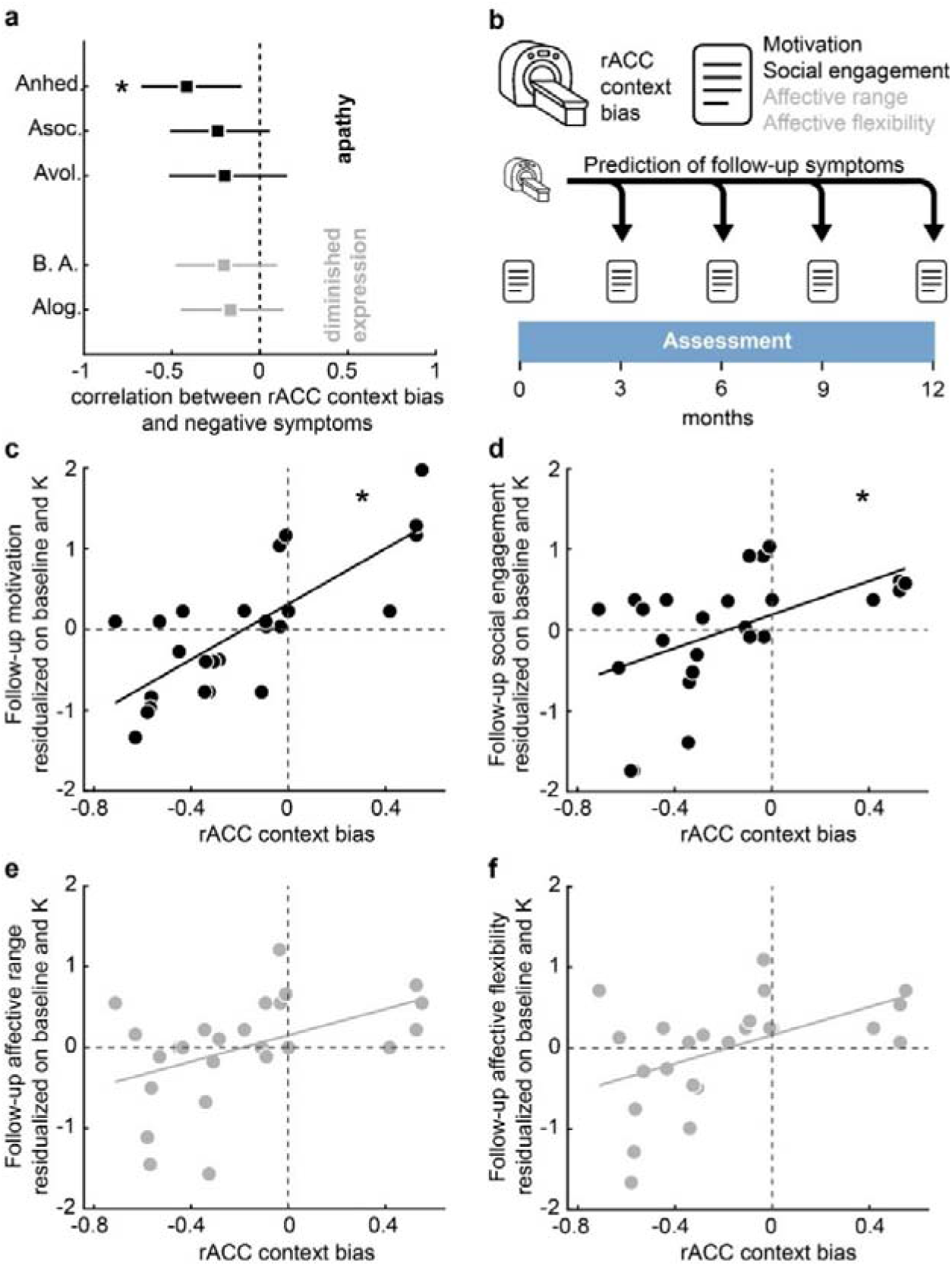
Right ACC context bias and negative-symptom outcomes. **(a)** Baseline associations: Correlations between the goal-directed context bias in the right anterior cingulate cortex (rACC) and BNSS domains. Black marks indicate the apathy cluster (anhedonia, asociality, avolition); gray marks indicate the diminished-expression cluster (blunted affect, alogia). Lines show 95% confidence intervals. **(b)** Schematic of the longitudinal analysis: rACC context bias measured at baseline (fMRI) was related to negative-symptom features derived from AMDP chart ratings at 3, 6, 9, and 12 months. For each feature, we summarized the average follow-up severity across the available assessments; K denotes the number of follow-up assessments for each participant. **(c–f)** Relationship between baseline goal-directed context bias in the rACC and follow-up motivation, social engagement, affective range, and affective flexibility. Points show follow-up scores adjusted for baseline and K, and lines show linear fits. In apathy-related domains, a more loss-biased rACC signal (more negative values on the x-axis) is associated with worse adjusted outcomes over the next year, with the strongest association for motivation. Abbreviations: Anhed.: Anhedonia; Asoc: Asociality; Avol.: Avolition; B.A.: Blunted affect; Alog.: Alogia. Asterisks indicate *P* < 0.05.

## Results

The SSD cohort and HC showed no significant differences in age, sex, or ethnicity (**Table 1**). However, participants with SSD had lower educational attainment scores compared to controls (SSD: 1.66 ± 1.17; controls: 2.23 ± 1.29, *p* = 0.02). All participants with SSD received antipsychotic medication. Symptom severity was mild to moderate (BPRS total score: 39.04 ± 10.43). Negative symptom ratings on the BNSS showed both apathy-related symptoms (anhedonia: 10.68 ± 4.63; avolition: 6.5 ± 2.76; asociality: 6.06 ± 2.65) and diminished-expression components (blunted affect: 9.5 ± 4.55; alogia: 4.34 ± 2.48).

**Table 1.** Participant characteristics. Clinical symptoms were assessed using the Brief Psychiatric Rating Scale (BPRS) for general psychopathology and the Brief Negative Symptom Scale (BNSS) for negative symptoms. Cognitive performance was evaluated with the MATRICS Consensus Cognitive Battery (MCCB), reported as standardized T-scores. Antipsychotic medication dosage is reported in chlorpromazine equivalents (CPZ eq.). Data are presented as mean (standard deviation) or count (percentage). Educational attainment is coded as follows: 0 = mandatory schooling, 1 = secondary education (high school or equivalent), 2 = vocational training, 3 = higher professional education, 4 = university degree. HC: Healthy controls. SSD: Schizophrenia spectrum disorders.

| Characteristic | HC | SSD | p-value |
| --- | --- | --- | --- |
| **Demographics** |  |  |  |
| Age (years), mean (SD) | 28.00 (5.57) | 27.14 (6.42) | 0.481 |
| Sex (% female) | 20 (41.7) | 11 (22.0) | 0.061 |
| Ethnicity, n (% Caucasian) | 38 (79.2) | 40 (80.0) | 0.995 |
| % Asian | 6 (12.5) | 6 (12.0) |  |
| % African | 4 (8.3) | 4 (8.0) |  |
| Educational attainment, mean (SD) | 2.23 (1.29) | 1.66 (1.17) | 0.024 |
| **Illness Characteristics** |  |  |  |
| Disease Duration (months), mean (SD) | NA | 18.26 (29.63) |  |
| Hospitalizations, mean (SD) | NA | 6.96 (4.98) |  |
| **Medication Status** |  |  |  |
| Dosage, antipsychotic (CPZ eq.) | NA | 277.20 (160.93) |  |
| **Clinical Ratings** |  |  |  |
| BPRS Total Score, mean (SD) | NA | 39.04 (10.43) |  |
| Suspicion | NA | 1.94 (1.10) |  |
| Hallucination | NA | 1.84 (1.13) |  |
| BNSS Total Score, mean (SD) | NA | 40.44 (15.37) |  |
| Asociality | NA | 6.06 (2.65) |  |
| Avolition | NA | 6.50 (2.76) |  |
| Anhedonia | NA | 10.68 (4.63) |  |
| Distress | NA | 3.32 (1.49) |  |
| Blunted Affect | NA | 9.50 (4.55) |  |
| Alogia | NA | 4.34 (2.48) |  |
| **Cognitive Performance** |  |  |  |
| MATRICES - Attention | NA | 0.97 (1.06) |  |
| MATRICES - Working Memory | NA | 1.47 (0.86) |  |
| MATRICES - Verbal Learning | NA | 1.59 (1.13) |  |
| MATRICES - Reasoning | NA | 1.44 (1.05) |  |
| MATRICES - Visual Learning | NA | 1.50 (0.90) |  |

### Decision behavior in reward and loss contexts

In the raw behavioral analysis (B_raw_, see Methods), individuals with SSD showed reduced model-based control compared to HC in both reward (p = 0.005) and loss contexts (p = 0.014; **Supplementary Fig. 1**). Within-group comparisons revealed that HCs exhibited a numerically higher contribution of model-based control in the reward relative to the loss context, whereas individuals with SSD showed the opposite pattern—a numerically higher contribution in the loss relative to the reward context (**Fig. 2a, Supplementary Fig. 5a**). However, these context-dependent differences were not statistically significant (p = 0.28).

To obtain a more nuanced estimate of model-based versus model-free contributions, we applied a hybrid reinforcement learning model. Consistent with the raw behavioral findings, the hybrid model confirmed reduced model-based control in individuals with SSD in the reward context (*p* = 7.9 × 10⁻¹³), but not in the loss context, when comparing individual-level ω parameters across groups (**Supplementary Fig. 5b**). The model further revealed a significant reward bias in HCs (*p* = 3.6 × 10⁻L) and a significant loss bias in the SSD group (*p* = 2.9 × 10⁻L). The difference in context-dependent biases between groups was significant (*p* = 2.8 × 10⁻¹L; **Fig. 2b**). We also examined whether ω-context bias was related to age, sex, illness duration, antipsychotic dose (chlorpromazine equivalents), or 5-HT_2A_ and D_2_ receptor occupancy (seeLMethods), but none of these associations reached significance. Across all contexts and groups, ω and B_raw_ were strongly correlated (*r*_s_ = 0.60–0.70, *p* ≤ 1 × 10⁻L in every case), indicating that B_raw_ reliably approximates the model-derived estimate and similarly captures the balance between model-based and model-free control. In the reward context, the posterior distribution of group mean differences confirmed that individuals with SSD had a lower model-based weight parameter (ω), as well as lower learning rates at both the first (α_1_) and second stage (α_2_), and reduced exploitation at the first stage (β_1_) compared to HC (HC minus SSD, 95% HDIs; Δω: 0.16 – 0.68; Δα1: 0.04 – 0.86; Δα2: 0.16 – 0.65; Δβ1: 0.23 – 3.29; **Supplementary Fig. 6**). In the loss context, the posterior distribution indicated significantly lower second-stage learning rates and reduced exploitation at both stages in the SSD group, while the model-based weight parameter (ω) did not differ between groups (HC minus SSD, 95% HDIs: Δα2: 0.31 – 0.9; Δβ1: 0.61 – 3.57; Δβ2: 0.06 – 1.37; **Supplementary Fig. 7**). Both in the reward and loss setting, MCMC samples of all model parameters were well-mixed and converged to stationary distributions (**Supplementary Figs 8 and 9)**. Convergence to stationary distributions was additionally indicated by the index of convergence (L across parameters: 1.00 to 1.04 in the reward and loss setting). Parameter-recovery analysis confirmed that all seven hybrid-model parameters were identifiable: across contexts, all Spearman correlations between true and recovered values of the seven parameters were positive and significant (*p*□≤□0.001 in every case; ω: r_s_ = 0.72 in the reward context; r_s_ = 0.76 in the loss context; for more details, see **Supplementary Figs 2-3**).

### Context-dependent neural activations

In the ACC, HC showed a significant reward bias in both hemispheres (*signrank test*; left and right: p = 0.001), while individuals with SSD exhibited a numerical loss bias. Group comparisons revealed a significant difference in context bias for both the left and right ACC (*ranksum test*; left and right: *p* = 6*10^-4^, **Fig. 3a, b**). In the DLPFC, a significant reward bias was found in HC in both hemispheres (*signrank test*; left: p = 0.002; right: p = 0.01). In the left hemisphere, this reward bias differed significantly from the numerical loss bias observed in individuals with SSD (ranksum test: p = 0.02, **Fig. 3a**). In the OFC, HC showed a significant reward bias in both hemispheres (*signrank test*; left: *p* = 0.001; right: *p* = 0.01), while individuals with SSD demonstrated a significant shift toward a numerical loss bias (*ranksum test*; left: *p* = 0.001; right: *p* = 0.002, **Fig. 3a, b**). Context-biased activation within ROIs was predominantly observed in central portions of the DLPFC and OFC in both groups. Within the ACC, individuals with SSD exhibited loss-biased activation predominantly in dorsal regions, whereas reward-biased activation in controls was stronger in ventral regions (**Fig. 3c, d**).

We next examined whether context bias in any of these regions was associated with negative symptom domains. Only the context bias in the right ACC showed a significant negative correlation with the symptom domain of anhedonia (*r*_s_ =-0.41; p = 0.048; **Fig. 4a**). Thus, greater loss-biased activation in rACC was associated with greater anhedonia. As an additional control, we regressed anhedonia on rACC context bias while adjusting for age, sex, illness duration and antipsychotic dose (see Methods); the bias remained a significant negative predictor (β□=□–4.26□±□1.51SE,□*p*□=□0.008). We also tested whether regional context bias correlated with age, sex, illness duration, antipsychotic dose (chlorpromazine equivalents), HT_2A_-and D_2_-occupancy according to their medications (see Methods); none of these associations reached significance. Apart from the aLpriori regions of interest (ACC, OFC, and DLPFC), our exploratory fMRI analysis identified additional areas showing reward-or loss-biased activation during model-based control (**Supplementary Fig. 10**). During model-based choices, individuals with SSD showed loss-biased activity centered in anterior and posterior cingulate as well as temporal regions, whereas reward-biased responses were observed mainly in occipito-temporal regions (**Supplementary Table 6**). By contrast, HC expressed a broad reward-biased network spanning for instance cingulate and paracingulate regions, parietal, orbitofrontal and supramarginal regions, thalamus, angular gyrus as well as further medial and dorsolateral prefrontal regions, with only a small occipital-pole cluster showing the opposite loss-dominant pattern (**Supplementary Table 7**).

### Baseline rACC context bias predicts follow-up negative-symptom–related outcomes

In the SSD subset with longitudinal clinical data (n=25), particularly baseline anterior cingulate context bias during goal-directed control was prospectively associated with negative-symptom-related outcomes over follow-up (**Fig. 4b-f**). In ANCOVA, greater loss bias in rACC predicted worse baseline-adjusted follow-up levels (mean 3–12 months) in several negative-symptom–related domains. The association was strongest for motivation (β = 1.83, 95% CI 1.20–2.46, *p* = 3.18×10^⁻5^; partial *R*² = 0.64; model *R*² = 0.79), and significant effects were also observed for social engagement (β = 1.11, 95% CI 0.31–1.92, *p* = 0.03; partial *R*² = 0.28; model *R*² = 0.54, **Fig. 4c, d**). A similar but smaller effect on motivation (β = 1.55, 95% CI 0.86–2.25, *p* = 4×10^-3^; partial *R^2^* = 0.51; model *R^2^* = 0.718) and a similar effect on social engagement (β = 1.15, 95% CI 0.4–1.91, *p* = 0.03; partial *R^2^* = 0.33, model *R^2^* = 0.57) was present in the lACC. In addition, the lOFC showed a weak yet significant association between context-biased activation and social engagement (β = 0.91, 95% CI 0.171–1.65, *p* = 0.04; partial *R^2^* = 0.238; model *R^2^* = 0.51). No other ROI showed a significant association with the course of negative-symptom–related domains, and no ROI was significantly related to the course of affective range or affective flexibility. The largest associations with affective range and affective flexibility were also observed in the rACC, positive in direction but did not survive multiple-comparison correction (**Fig. 4e, f**). In summary, the longitudinal analysis indicates that greater baseline loss bias, particularly in anterior cingulate regions, forecasts poorer subsequent apathy-related outcomes, most prominently in motivation.

## Discussion

The present study provides converging behavioral and neural evidence that SSD are characterized by a systematic re-allocation of goal-directed cognitive resources away from reward pursuit and toward loss avoidance. Relative to the reward-biased deployment of model-based strategies in HC, individuals with SSD showed model-based control primarily when potential losses were at stake. This loss-biased strategy was mirrored at the neural level by preferential engagement of the bilateral ACC, OFC and left DLPFC during model-based learning in the loss condition. Only the degree of loss-biased ACC activation tracked the severity of a negative symptom domain at baseline, given the association with anhedonia. Critically, baseline loss-biased engagement in the right anterior cingulate cortex also carried prognostic information: A more loss-biased ACC context-bias signal predicted poorer follow-up levels in several negative-symptom–related domains, with the strongest effect for motivation. Together, these results offer a mechanistic explanation for the clinical observation that individuals with schizophrenia show impaired pursuit of positive outcomes yet intact loss avoidance^5, 6^, while linking this asymmetry to the negative symptom domain anhedonia and to the subsequent course of negative symptom-related outcomes. It is worth mentioning that the raw behavioral analysis showed only a non-significant numerical context effect, while the hierarchical reinforcement learning model revealed a robust difference. In contrast to the raw behavioral analysis, the hierarchical reinforcement-learning model fits the full choice history and jointly estimates learning rate, perseveration, and lapse parameters, and is therefore better suited to distill the goal-directed context effect. Hierarchical pooling further reduces within-group noise through shrinkage, increasing power to detect between-group differences^51^. Given these analytic differences, an underlying context effect that is subtle in the coarse, previous-trial-based summary analysis of raw behavior is likely to emerge more robustly when estimated with the full hierarchical hybrid model.

Canonical reinforcement-learning theory assumes that model-based control is deployed when the expected benefits of flexible planning outweigh its cognitive costs^4, 9^. In healthy adults, appetitive contexts typically enhance the perceived value of planning, leading to robust model-based control in reward contexts, as observed in our control sample and in previous studies^44, 46^. The loss-biased pattern in participants with SSD suggests that for these individuals the subjective cost–benefit calculus is distorted: avoiding losses is deemed more valuable than obtaining gains. One parsimonious interpretation is that patients discount prospective rewards because they anticipate diminished pleasure from realizing potential gains^57^, whereas losses remain fully salient. In line with this idea, low “decision acuity”—a trait-like reduction in overall planning capacity marked by noisier, short-sighted choices^58^— could further restrict cognitive resources and channel them chiefly into preventing losses, thereby producing the loss-biased goal-directed control we observe. A complementary line of evidence shows that people with SSD are less willing to expend cognitive effort even for large rewards, pointing to a domain-general undervaluation of appetitive incentives^59, 60^. These findings align with evidence for impaired value representation during reward processing relative to preserved loss avoidance in SSD^5^, and our results frame this as the consequence of a strategic shift toward loss-oriented cognitive control.

Our fMRI results reveal how the cognitive bias towards loss avoidance in individuals with SSD is implemented in the prefrontal cognitive network. In controls, reward-related increases in model-based control were accompanied by greater activation of bilateral ACC, OFC and left DLPFC—regions known to orchestrate cost-dependent allocation of cognitive resources, value representation, and prospective planning according to goal-representation^4, 47^. In individuals with SSD, the same circuit was preferentially engaged for model-based control when potential losses, rather than rewards, were at stake. These findings extend prior observations of blunted OFC and DLPFC responses to positive outcomes^8^ by demonstrating that computations underpinning flexible decision-making are context-dependent and retuned toward loss avoidance in psychotic disorders. While loss-dominated activation in the OFC of individuals with SSD may signal a heightened valuation of avoiding negative outcomes, a similar activation pattern in the left DLPFC might reflect increased recruitment of cognitive resources during the planning of loss avoidance compared to reward seeking^4, 42, 43, 46^. The ACC emerged as a particularly pivotal hub: its loss-biased engagement was the only neural measure that covaried with a negative symptom domain. The ACC is ideally positioned to integrate motivational signals with task-control demands and to evaluate whether engaging the additional cognitive effort required for model-based learning is worthwhile^40, 42, 43^. A loss-dominated encoding of action value within the ACC could therefore funnel limited cognitive resources into preventing negative outcomes, leaving insufficient capacity for goal-directed reward processing and manifesting clinically as anhedonia. Our longitudinal analysis suggests that this anterior cingulate control signature is not merely epiphenomenal but captures variance in later negative-symptom-related outcomes beyond baseline levels, consistent with a candidate neurocomputational marker for risk stratification. Interestingly, the association between rACC loss bias and current as well as future negative-symptom– related outcomes was evident for apathy-related domains but not for diminished-expression-related components. This pattern is consistent with accounts positing a link between aberrant valence processing and negative symptoms specifically within the apathy dimension^48, 61^, and it further specifies an rACC-centered goal-directed control mechanism with prognostic relevance.

Our findings converge with mounting evidence that the ACC is central to psychotic disorders, with ACC dysfunction contributing to impairments in proactive control, and uniform structural features characterizing this region in schizophrenia^4, 41, 62^. Identifying a loss-biased planning mode in schizophrenia has potential translational relevance. First, it provides a neurocomputational phenotype that could be quantified and used to stratify patients or monitor treatment responses. Second, the association between anhedonia and ACC loss bias positions the anterior cingulate cortex as a promising target for circuit-based interventions. Non-invasive brain stimulation targeting the ACC or novel pharmacological interventions may re-balance resource allocation and thereby ameliorate motivational deficits. Our findings also extend earlier active-inference results showing elevated forgetting and reduced policy precision during action selection in early psychosis^63^, and place the concomitant computational profile in a motivational framework that distinguishes loss from reward contexts.

This study has several limitations. First, our neurocomputational findings are cross-sectional and cannot establish whether loss-biased planning predates illness onset, emerges progressively or reflects a secondary adaptation to chronic disability. Longitudinal studies in high-risk and first-episode cohorts are needed to pinpoint the emergence of loss-biased cognitive processing in schizophrenia. Second, participants were medicated, and systematic modulation of cognitive processing and behavioral strategies by the complex receptor modulation of antipsychotic agents^56, 64, 65^ cannot be excluded. However, we found no associations between context-dependent brain activation or behavior and chlorpromazine equivalents, 5-HT2A and D2 receptor occupancy. These findings argue against the interpretation that loss-biased behavior and brain activation observed in individuals with SSD merely reflect medication effects. Third, a recent study showed that modeling parameters in reinforcement tasks can vary over time, potentially limiting their capacity for precision psychiatry^66^. In our case, a within-subject contrast of reward and loss context may attenuate common-mode fluctuations such as global response scaling and lapse-related noise, thereby yielding greater longitudinal stability than single-context parameters. Finally, the monetary outcomes used here do not capture the rich spectrum of rewards and losses encountered in daily life, and virtual-reality tasks embedding social, vocational and other incentives could enhance ecological validity.

In summary, by integrating computational modeling with neuroimaging, we demonstrate that SSD are not marked by a uniform deficit in goal-directed cognition but by a selective redirection of cognitive resources toward loss avoidance. This behavioral shift is instantiated in a prefrontal network comprising ACC, OFC and DLPFC, and loss-biased engagement of the ACC is coupled to the negative symptom domain anhedonia. Our findings provide a mechanistic explanation for the preservation of avoidance learning alongside impaired reward pursuit in schizophrenia and outline circuit-level targets for interventions aimed at restoring adaptive motivation.

## Data Availability

All data produced in the present study are available upon reasonable request to the authors.

## Acknowledgements

The authors thank Dr.LFranziska Knolle for her critical reading of the manuscript and her valuable scientific advice.

## Author Contributions

W.O. and P.H. designed the experiments. G.C. and W.O. performed the experiments. W.O., A.M., G.H. and G.C. analyzed the data. X.W., V.E., G.P., B.Q., P.T. contributed to the concept of the data analysis and manuscript. P.H. initiated and supervised the study. All authors approved, refined and edited the final version of the manuscript.

## Data and Code availability

Upon publication, all de-identified data underlying the results and the analysis code will be deposited in a public repository with persistent identifiers (DOIs) and an open license; repository links will be added to the final article.

