## Supplementary Material for "Goal-Directed Control in Schizophrenia: Loss-Biased Engagement of the Anterior Cingulate Relates to Negative-Symptom Outcomes"

***Computational Modeling***

In the two-stage decision task, participants navigate through a sequence of states and actions designed to capture two complementary learning systems Model-based (MB) and Model-free (MF) learning. The set up includes three states: a first-stage state ($s_{1}=s_{A}$) and two possible second-stage states ($s_{2}=s_{B}$ or $s_{2}=s_{C}$), with each state associated with two actions ($a_{A}$ and $a_{B}$). At the start of each trial $t$, participants are always at state $s_{A}$, The transition to one of the two second-stage states ($s_{2,t}$) depends probabilistically on the action taken at the first stage ($a_{1,t}$). Reward ($r_{2,t}$), is only received at the end of the second stage (which can be 0 or 1 in the reward setting, and 0 or -1 in the loss setting), no reward is given at the end of first stage ($r_{1,t}$ = 0). During this task learning in the first stage is denoted as $\alpha_{1}$, while in the second stage as $\alpha_{2}$, eligibility trace ($\lambda$) captures how much an agent considers past rewards in updating the future actions, while the discount factor ($\gamma$) captures the importance of future rewards in updating the state-value function.

During the task, participants update their choices by learning a state-action value function $Q_{Net}$, which represents the expected future reward of taking action $a$ in state $s.$This state value function can be further decomposed into two components, a model-free and a model-based component.

At the start of each trial $t$ and stage $i$, the model-free component $Q_{MF}$,is updated as:

$Q_{MF}(s_{i,t},a_{i+1,t})\leftarrow Q_{MF}(s_{i,t},a_{i,t})+\alpha_{i}\delta_{i,t}$	[1]

where Reward Prediction Error (RPE $\delta_{i,t}$) is defined as:

$\delta_{i,t}=r_{i,t}+Q_{MF}\left( s_{i+1,t},a_{i+1,t} \right)-Q_{MF}\left( s_{i,t},a_{i+1,t} \right)$	[2]

At the start of the trial all state-action values are initialized to zero before learning begins, meaning $Q_{MF}\left( s_{2,1},a_{2,1} \right)=Q_{MF}\left( s_{1,1},a_{1,1} \right)= 0$.

At trial $t=1$, we can illustrate how the learning process begins. Reward Prediction Error (RPE) for the second stage $i=2$ is calculated based on eq(2) :$\delta_{2,1}=r_{2,1}-Q(s_{2,1},a_{2,1})$. Since there is no third stage, the value function for any future state $Q_{MF}(s_{3,t},a_{3,t})$ is always defined to be zero. Consequently, the RPE for the second stage on the very first trial is simply equal to the reward received $r_{2,1}$. This initial RPE is then used to update the value of the chosen second-stage action. Following the update rule from Equation 1, the new value function for stage 2, becomes $Q_{MF}(s_{2,1},a_{2,1})\leftarrow0+\alpha_{2}\delta_{2,1}$. Now, for stage 1, as there is no reward $r_{1,t}$ is always 0, the RPE becomes from eq (2): $\delta_{1,1}=Q(s_{2,1},a_{2,1})$. Additionally, the eligibility parameter (λ) (Daw 2011), controls the degree to which the outcome of the second stage influences the learned value of the initial first-stage choice eq(3)

$Q_{MF}(s_{1,t},a_{1,t})\leftarrow Q_{MF}(s_{1,t},a_{1,t})+\alpha_{1}\lambda\delta_{2,t}$	[3]

This process is iterated across all the trials to calculate how MF RPE evolves for each participant during the task.

In contrast, the model-based system operates by first constructing an internal model, or "map," of the environment. This model contains the learned probabilities of transitioning between states and the rewards available at each one. The system leverages this internal map to prospectively compute an action's value by planning through potential outcomes.

Applying this to the current task, a model-based strategy first requires learning which first-stage choices are likely to lead to specific second-stage states. It then uses this knowledge to "plan ahead" by assessing the rewards available in those future states. Since the first stage never yields a reward, the value of an initial action is determined entirely by the anticipated rewards of the second stage it is expected to reach.

In our current setup, we have the following transition probabilities $P(s_{B}|s_{A},a_{A})=P(s_{C}|s_{A},a_{B})=0.7$ and $P(s_{C}|s_{A},a_{A})=P(s_{B}|s_{A},a_{B})=1-0.7=0.3$.

At the second stage, since there are no further stages to anticipate, the state-value function for MB learning is the same as for MF learning.

$Q_{MB}(s_{2,t},a_{2,t})=Q_{MF}(s_{2,t},a_{2,t})$	[4]

While for the second stage as, the model-based algorithm takes into consideration the transition probabilities and tries to maximize the value it is calculated as:

$Q_{MB}(s_{A,t},a_{A,t})=\sum_{s_{2}\in B,C} P(s_{2}\mid s_{A,t},a_{A,t})\max_{a_{2}\in a_{A},a_{B}}Q_{MB}(s_{2,t},a_{2,t})$	[5]

These values are also updated for each trial iteratively. Now, as learning is driven by both MF and MB process, the learned state value functions are a combination eq(6) and are controlled using a weighting parameter $\omega$. For a complete MB system $\omega=1$ while for a full MF system it is $\omega=0$.

Now, the combined value in the first stage becomes:

$Q_{Net}=\omega Q_{MB}\left( s_{A},a_{j} \right)+\left( 1-\omega\right)\left( s_{A},a_{j} \right)$	[6]

Whereas, at the second stage, $Q_{net}=Q_{MB}=Q_{MF}$. Finally, to convert the state-value functions into action probabilities to represent choices, we use a softmax function:

$P(a_{i,t}=a|s_{i,t})=\frac{exp(\beta_{i}[Q_{net}(s_{i,t},a)+p\cdot rep(a)])}{\sum_{a\mathcal{\in A}} exp(\beta_{i}[Q_{net}(s_{i,t},a^{'})+p\cdot rep(a^{'})])}$	[7]

where, $\mathcal{A}$ represents the set of actions at that state; the $\beta$ parameter controls how deterministic the choices are with a higher $\beta$ revealing exploitation and a lower $\beta$ value denoting more random choices; an exploratory behavior, $p$ represents the choice preservation for the first stage; and $rep(a)$ is an indicator function which indicator function that equals 1 if the previous first-stage choice was the same.

**MRI processing**

**fMRI scanning protocol**

Two consecutive fMRI scans were acquired. During the first (resting state), participants were instructed to relax and keep their eyes open. In the second, participants performed the modified version of the two-stage task in a “reward” and in a “loss” setting. Participants were monitored to ensure they remained awake and relaxed. They wore a respiratory belt to track breathing and were observed with a camera. Wakefulness was monitored throughout the scan, and customized head molds were used to reduce head motion. In addition, participants were provided with earplugs and headphones.

MRI was conducted on a Philips Achieva 3.0 T magnetic resonance scanner with a 32-channel SENSE head coil. During the resting state sequence, 900 echo-planar imaging volumes were recorded (TR = 2000 ms, TE = 30 ms, matrix = 64 x 64, FOV = 240 mm, slice thickness = 3 mm, 40 continuous axial oblique slices). Each session began with the acquisition of structural images, including a sagittal localizer, a high-resolution T1-weighted SPGR 3D spoiled gradient sequence (TR = 7.5 ms, TE = 3 ms, matrix = 256 × 256, FOV = 240 mm), and a T2 FSE sequence (TR = 6434 ms, TE = 102 ms, matrix = 256 × 256, slice thickness = 2.5 mm).

**Anatomical data processing**

The T1-weighted image was corrected for intensity non-uniformity (INU) with N4BiasFieldCorrection ^1^, distributed with ANTs 2.5.0 ^2^, and used as T1-weighted reference throughout the workflow. The T1-weighted reference was then skull-stripped with a Nipype implementation of the antsBrainExtraction.sh workflow (from ANTs), using OASIS30ANTs as the target template. Brain tissue segmentation of cerebrospinal fluid (CSF), white-matter (WM), and gray-matter (GM) was performed on the brain-extracted T1-weighted image using fast ^3^. Volume-based spatial normalization to one standard space (MNI152NLin2009cAsym) was performed through nonlinear registration with antsRegistration (ANTs 2.5.0) ^2^, using brain-extracted versions of both the T1-weighted reference and the T1-weighted template. The ICBM 152 Nonlinear Asymmetrical template version 2009c ^4^ was selected for spatial normalization and accessed with TemplateFlow.

**Functional data processing**

There was one BOLD run per subject. For pre-processing, a reference volume was first generated, using a custom methodology of *fMRIPrep*, for use in head motion correction. Head-motion parameters with respect to the BOLD reference (transformation matrices, and six corresponding rotation and translation parameters) were estimated before any spatiotemporal filtering using mcflirt ^5^. The BOLD reference was then co-registered to the T1-weighted reference using mri_coreg (FreeSurfer) followed by flirt with the boundary-based registration cost-function. Co-registration was configured with six degrees of freedom. Several confounding time series were calculated based on the *preprocessed BOLD*: framewise displacement (FD), DVARS, and three region-wise global signals. FD was computed following Power, Mitra, Laumann, Snyder, Schlaggar, Petersen ^6^ and Jenkinson, Bannister, Brady, Smith ^5^. FD and DVARS were calculated for each functional run, both using their implementations in *Nipype* ^6^.

The three global signals were extracted within the CSF, the WM, and the whole-brain masks. Additionally, a set of physiological regressors were extracted to allow component-based noise correction ^7^. Principal components were estimated after high-pass filtering the *preprocessed BOLD* time-series (using a discrete cosine filter with 128s cut-off) for the two *CompCor* variants: temporal (tCompCor) and anatomical (aCompCor). tCompCor components were then calculated from the top 2% variable voxels within the brain mask. For aCompCor, three probabilistic masks (CSF, WM and combined CSF + WM) were generated in anatomical space. The implementation differed from that of Behzadi, Restom, Liau, Liu ^7^ in that instead of eroding the masks by 2 pixels on BOLD space, a mask of pixels that likely contain a volume fraction of GM was subtracted from the aCompCor masks. This mask was obtained by thresholding the corresponding partial volume map at 0.05, preventing extraction from voxels with a minimal fraction of GM. Finally, these masks were resampled into BOLD space and binarized by thresholding at 0.99 (as in the original implementation).

Components were also calculated separately within the WM and CSF masks. For each CompCor decomposition, the *k* components with the largest singular values were retained, such that the retained components’ time series were sufficient to explain 50 percent of variance across the nuisance mask (CSF, WM, combined, or temporal). The remaining components were dropped from consideration. The head-motion estimates calculated in the correction step were also placed within the corresponding confounds file. The confound time series derived from head motion estimates and global signals were expanded with the inclusion of temporal derivatives and quadratic terms for each^8^. Frames that exceeded a threshold of 0.5mm FD or 1.5 standardized DVARS were annotated as motion outliers. Additional nuisance time series were calculated by means of principal components analysis of the signal found within a thin band (*crown*) of voxels around the edge of the brain.

All resamplings could be performed with *a single interpolation step* by composing all the pertinent transformations (i.e. head-motion transform matrices, susceptibility distortion correction when available, and co-registrations to anatomical and output spaces). Gridded (volumetric) resamplings were performed using nitransforms, configured with cubic B-spline interpolation. After images were preprocessed with fMRIPrep, preprocessed images were smoothed using a Gaussian kernel with full width at half-maximum of 8mm and were high-pass filtered with a cutoff of 128s. Four participants with SSD were omitted from further fMRI analysis: two owing to excessive head motion during fMRI acquisition, and two because scanner malfunction produced incomplete imaging data.

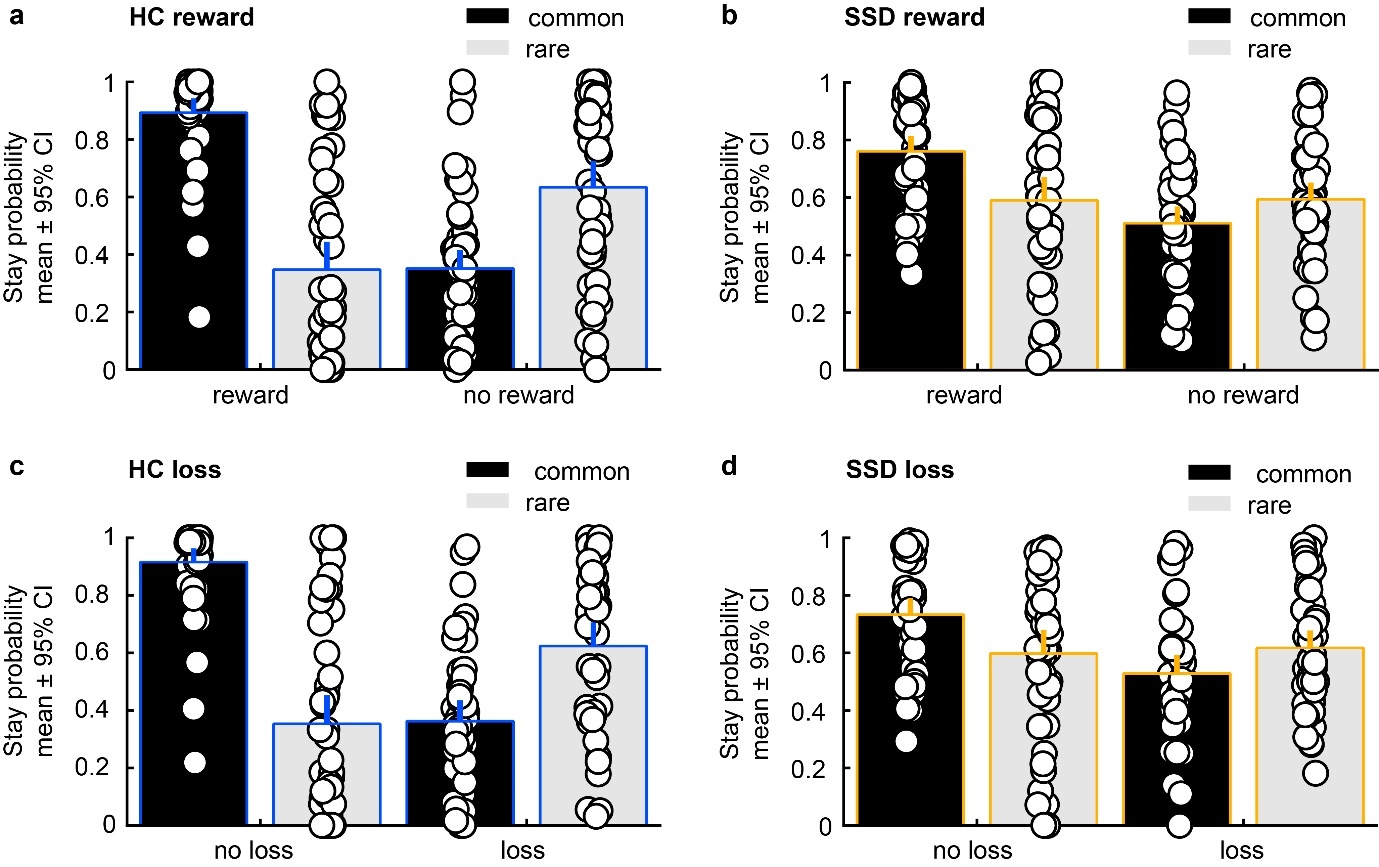

**Supplementary Figure 1: Analysis of the 2-stage decision task at the behavioral level.**
Left panel: Stay probabilities averaged across healthy controls (HC) in the reward (upper panel) and loss (lower panel) contexts (**a, c**). In both contexts, HC exhibit behavioral signatures consistent with both model-free and model-based learning. Right panel: Stay probabilities averaged across individuals with schizophrenia spectrum disorders (SSD) in the reward (upper panel) and loss (lower panel) contexts (**c, d**). Similar to HC, individuals with SSD show evidence of both model-free and model-based learning. Bars indicate group means ± 95 % confidence intervals; individual data points are overlaid.

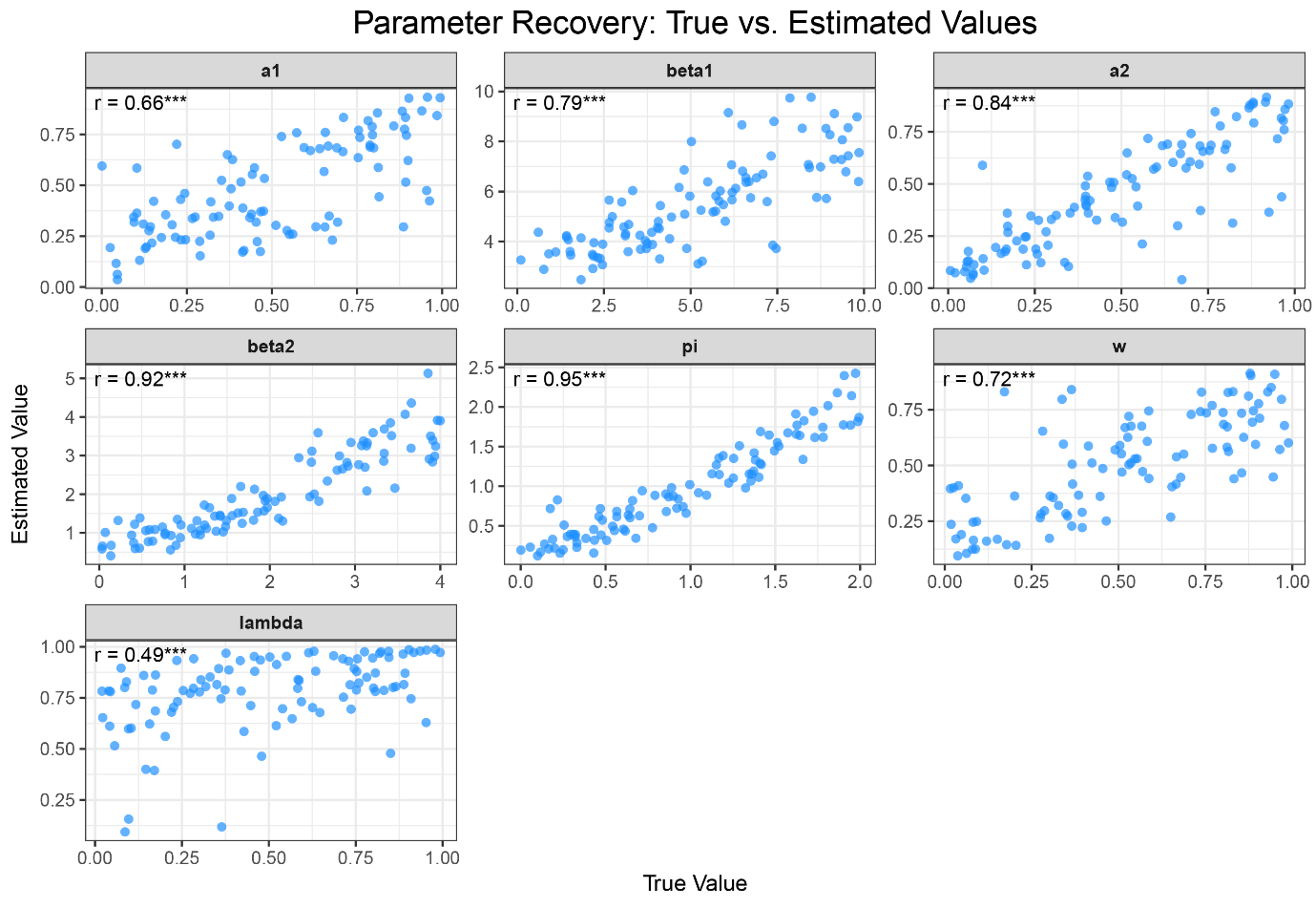

**Supplementary Figure 2: Parameter recovery (reward context) for the seven‑parameter two‑stage model.** For 100 simulated agents, we generated choice data from parameter sets and re‑fit the same model at the individual level. Each panel plots the recovered value against the ground‑truth generating value for one parameter—first‑stage learning rate (α₁), second‑stage learning rate (α₂), first‑stage inverse temperature (β₁), second‑stage inverse temperature (β₂), eligibility trace (λ), perseveration bias (π), and model‑based weight (ω). Points are simulated agents (n = 100). Annotations report Spearman’s ρ (true vs. recovered across agents); each agent completed 200 trials to match the empirical reward context block. *** denotes *p* < 0.001.

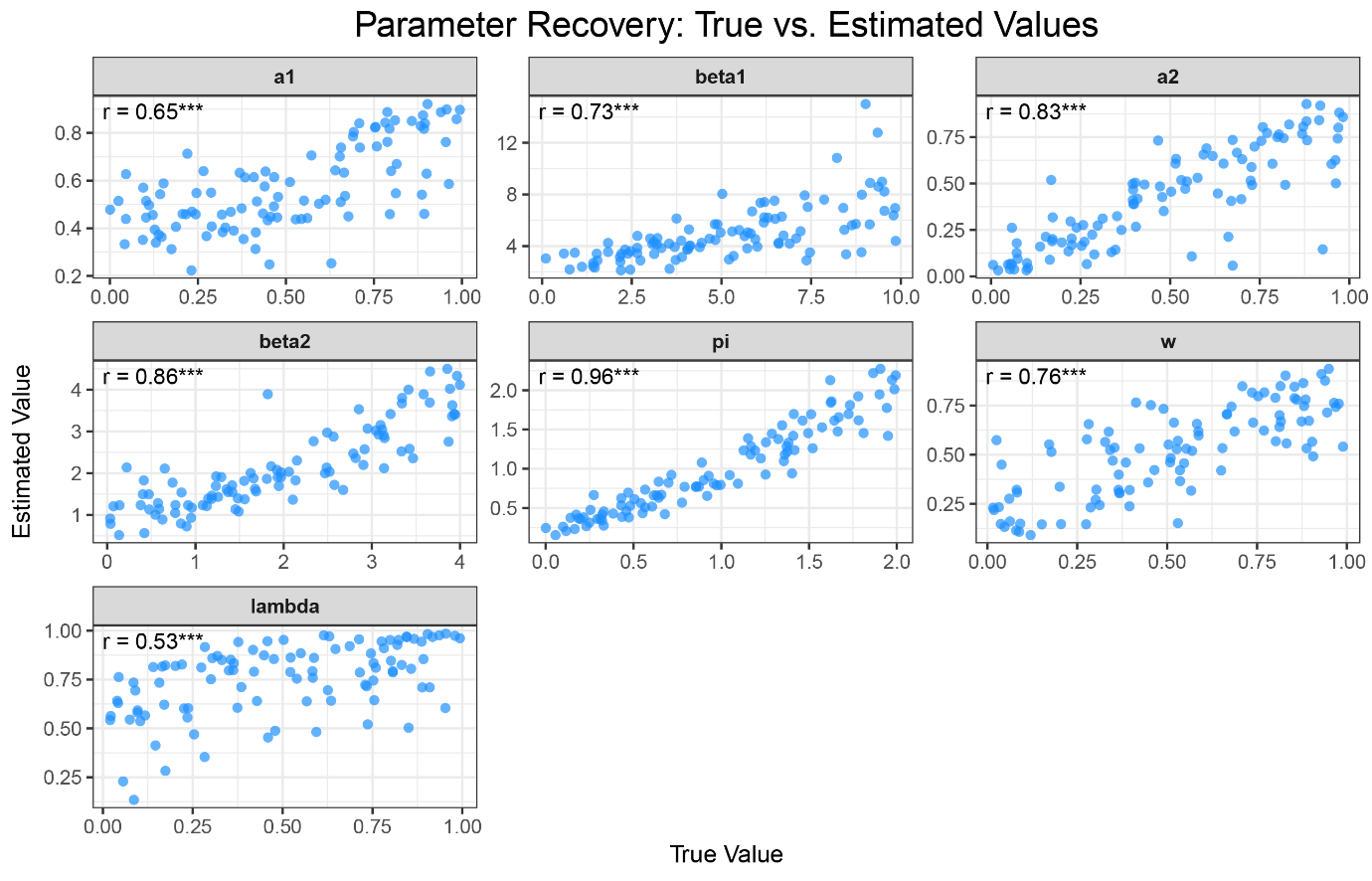

**Supplementary Figure 3: Parameter recovery (loss context) for the seven-parameter two-stage model.** Layout, axes, and statistical annotations are identical to those in Supplementary Fig. 2; each agent completed 200 trials to match the empirical loss context block.

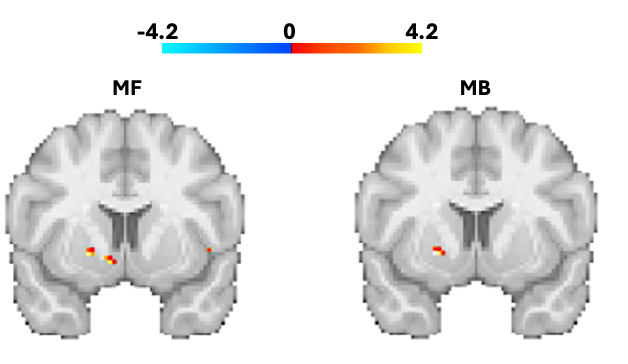

**Supplementary Figure 4:** **Neural correlates of model-free (left) and model-based (right) reward prediction errors in ventral striatum.** Similar, to Daw et al (2011)^9^, we also find activation in ventral striatum when healthy controls show model-based (MB) and model-free (MF) learning strategies in the reward context. Maps are thresholded for false-positive errors (0.005).

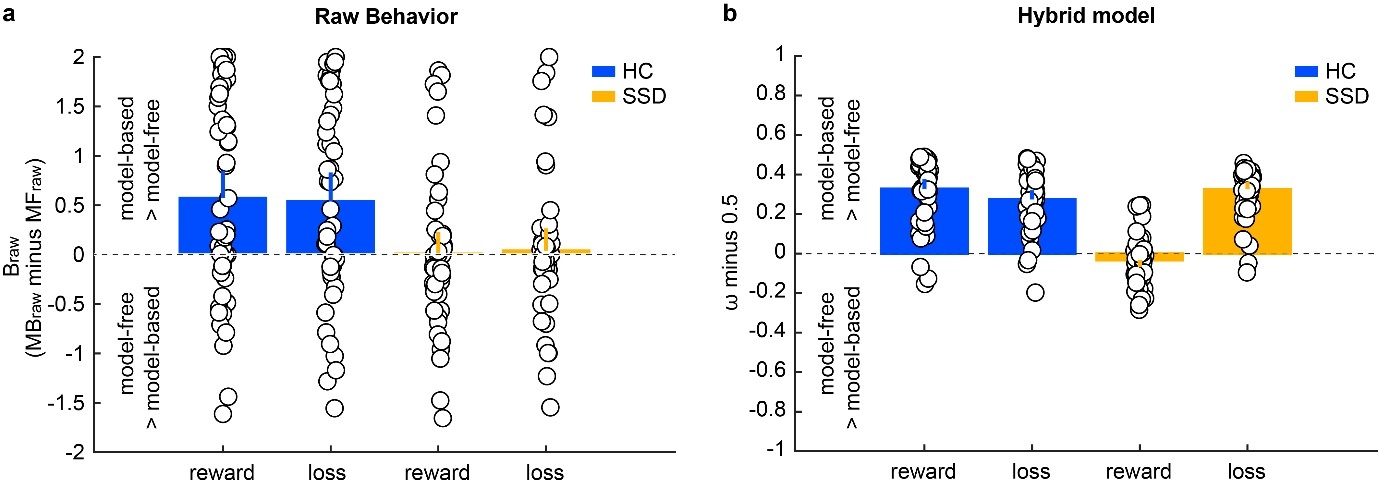

**Supplementary Figure 5: Raw and hybrid-model indices of model-based versus model-free control in reward and loss contexts.** (**a**) Raw behavioral scores for healthy controls (HC; blue) and individuals with schizophrenia-spectrum disorders (SSD; orange). Values > 0 indicate a predominance of model-based control; values < 0 indicate a predominance of model-free control. (**b**) Hierarchical hybrid-model scores (ω – 0.5) plotted on the same centered scale: 0.5 was subtracted from the ω-scores to facilitate comparison with the raw behavioral scores in (a); positive values reflect greater model-based, and negative values greater model-free influence. Bars show group means ± 95 % confidence intervals; individual data points are overlaid.

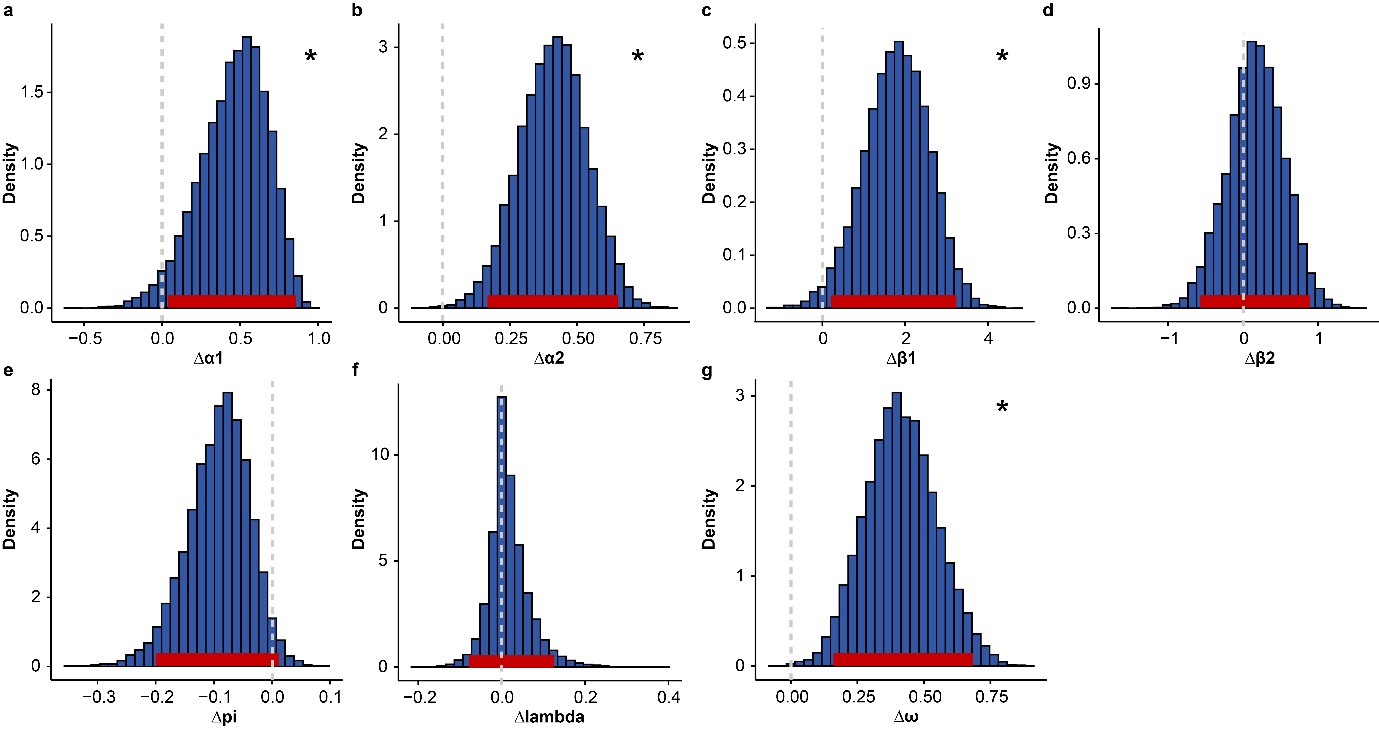

**Supplementary Figure 6:** **Group comparison of modeling parameters in the reward context.** (**a**) The posterior distribution of group mean differences (healthy controls (HC) minus schizophrenia-spectrum disorders (SSD)) is shown for the modeling hyperparameter α_1_. The red line indicates the 95% highest density interval (HDI). (**b-f**) Corresponding posterior distributions of group mean differences and 95%-HDIs are shown for the hyperparameters α_2_, β_1_, β_2_, pi, lambda and ω. Asterisks mark parameters whose 95 % highest‑density interval (HDI) excludes zero (significant group differences).

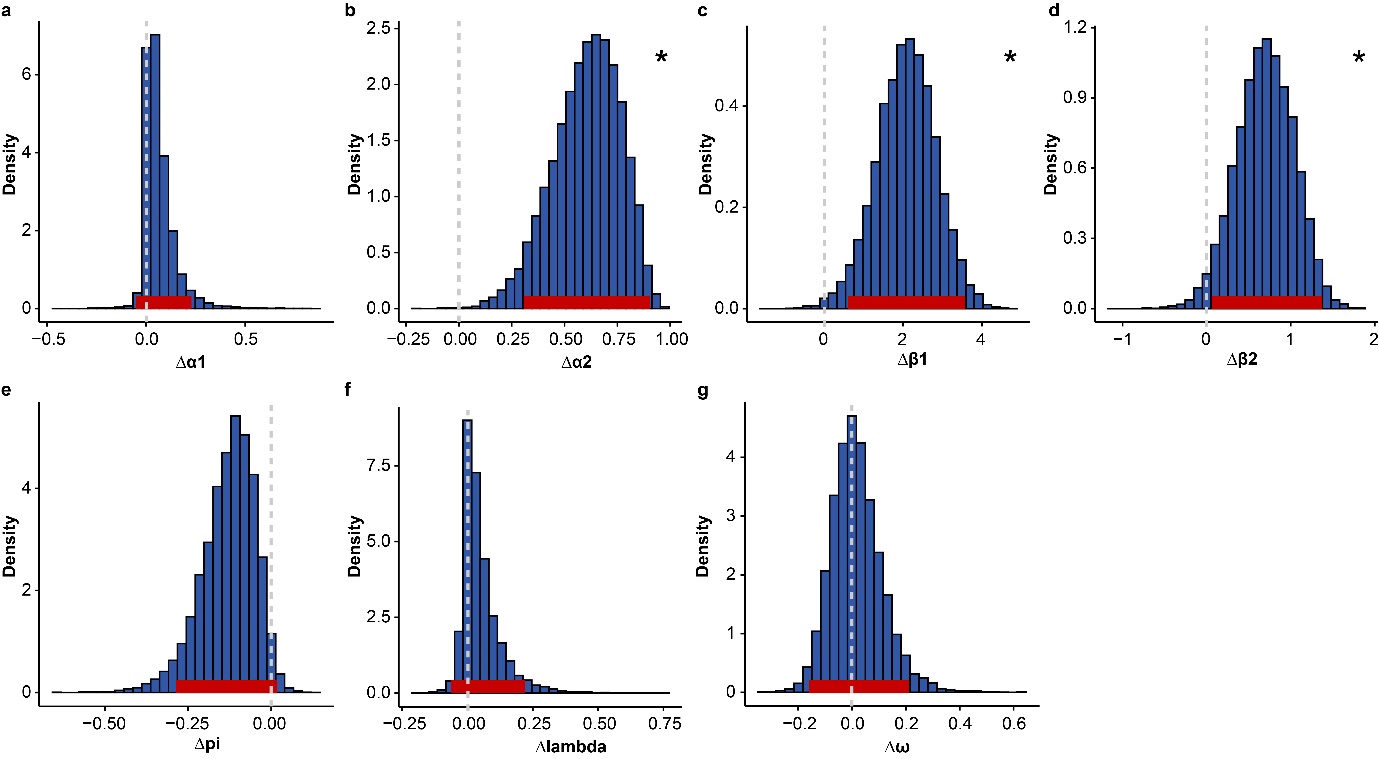

**Supplementary Figure 7:** **Group comparison of modeling parameters in the loss context. (a**) The posterior distribution of group mean differences (healthy controls (HC) minus schizophrenia-spectrum disorders (SSD)) is shown for the modeling hyperparameter α1. The red line indicates the 95% highest density interval (HDI). (**b-f**) Corresponding posterior distributions of group mean differences and 95%-HDIs are shown for the hyperparameters α2, β1, β2, pi, lambda and ω. Asterisks mark parameters whose 95 % highest‑density interval (HDI) excludes zero (significant group differences).

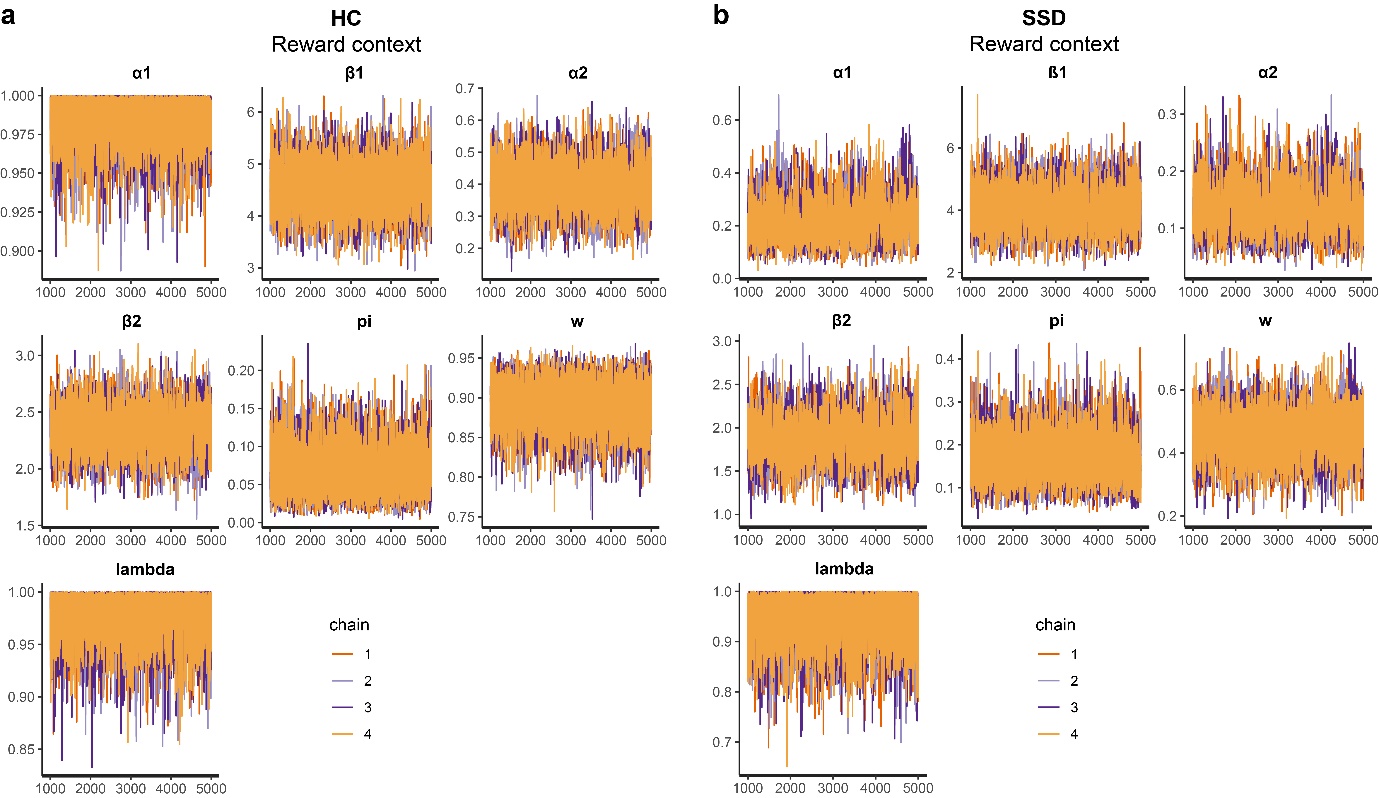

**Supplementary Figure 8:** **Markov Chain Monte Carlo (MCMC) sampling performance across modeling parameters in the reward condition.** (**a**) MCMC performance for the modeling hyperparameter α1 is shown based on 4000 post-burn-in samples and 4 chains. (**b-g**) MCMC performance is illustrated for the hyperparameters β_1_, α_2_, β_2_, pi, ω and lambda. Same conventions as in a. For all modeling parameters in the reward condition, MCMC samples are well-mixed and converged to stationary distributions.

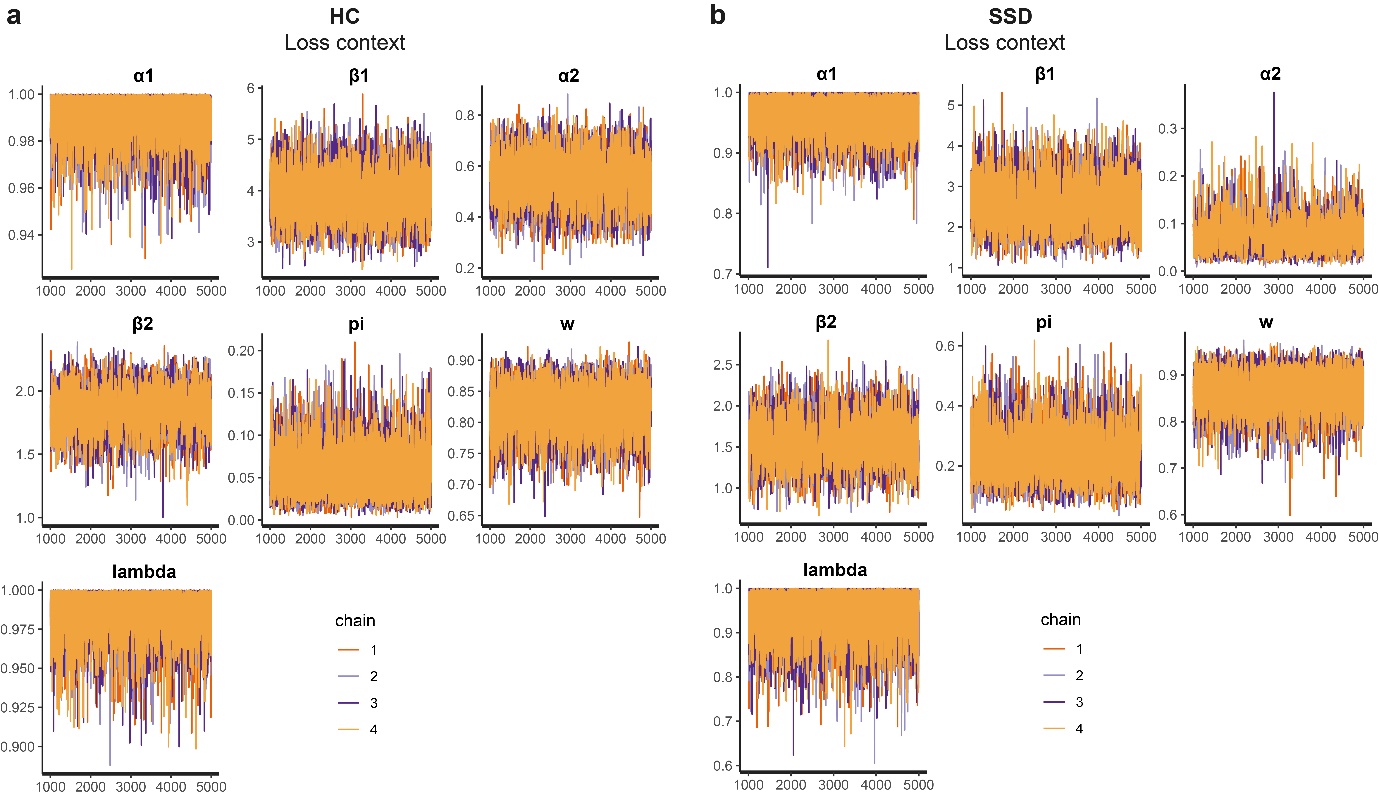

**Supplementary Figure 9:** **Markov Chain Monte Carlo (MCMC) sampling performance across modeling parameters in the loss condition.** (**a**) MCMC performance for the modeling hyperparameter α1 is illustrated based on 4000 post-burn-in samples and 4 chains. (**b-g**) MCMC performance is shown for the hyperparameters β_1_, α_2_, β_2_, pi, ω and lambda. Same conventions as in a. In the loss condition, the MCMC samples of all modelling parameters are well-mixed and converged to stationary distributions.

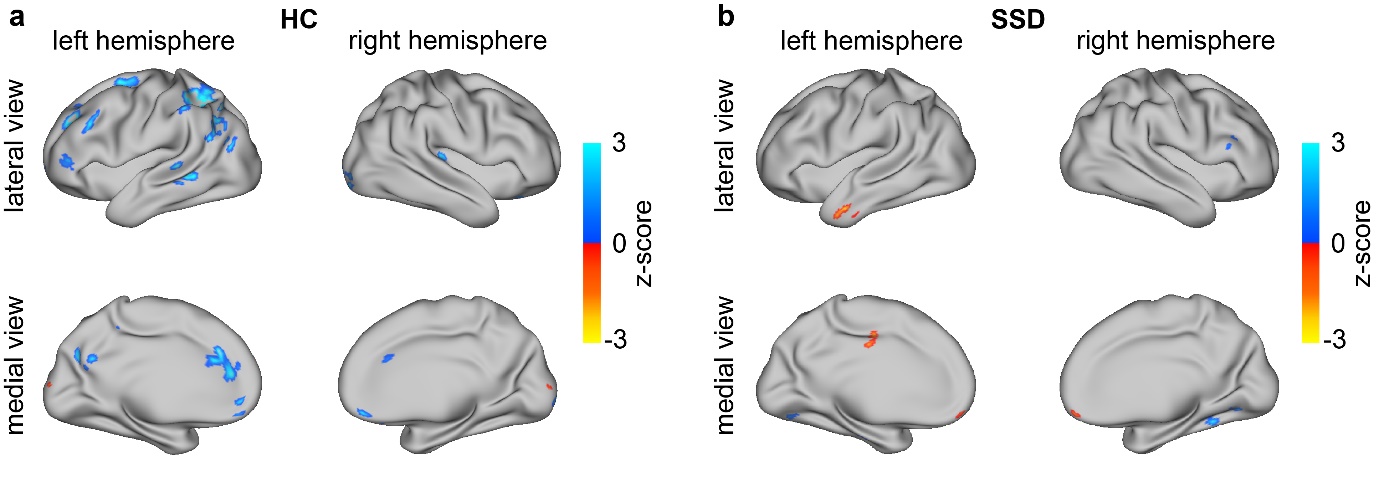

**Supplementary Figure 10: Whole-brain analysis reveals dissociable valence effects on model-based learning networks in individuals with schizophrenia spectrum disorders and healthy controls.** (a) Surface projections of significant z-scores are displayed for healthy controls (HC). Warm colors indicate brain regions with greater model-based signals for loss outcomes compared to reward outcomes, while cool colors indicate the reverse pattern. For visualization, Z-scores are projected onto the pial surface of a template brain. (**b**) Same for individuals with schizophrenia spectrum disorders (SSD).

| **Subject ID** | **0 months** | **3 months** | **6 months** | **9 months** | **12 months** |
| --- | --- | --- | --- | --- | --- |
| 5 | 1 | 1 | NA | NA | NA |
| 8 | 0 | 0 | NA | 0 | NA |
| 9 | 0 | 0 | NA | 0 | NA |
| 10 | 1 | 0 | NA | NA | NA |
| 11 | 0 | NA | NA | 1 | 0 |
| 12 | 0 | NA | NA | 0 | 2 |
| 13 | NA | NA | NA | NA | NA |
| 14 | NA | NA | NA | NA | NA |
| 15 | NA | NA | NA | NA | NA |
| 16 | NA | NA | NA | NA | NA |
| 17 | 1 | 2 | 0 | NA | 3 |
| 19 | NA | NA | NA | NA | NA |
| 20 | NA | NA | NA | NA | NA |
| 21 | 2 | 3 | 2 | NA | NA |
| 22 | 2 | 3 | NA | NA | 2 |
| 23 | 1 | 0 | NA | 0 | NA |
| 24 | 1 | 0 | 1 | 0 | 0 |
| 25 | NA | NA | NA | NA | NA |
| 26 | NA | NA | NA | NA | NA |
| 27 | NA | NA | NA | NA | NA |
| 28 | 1 | NA | 2 | NA | 2 |
| 29 | 1 | 2 | NA | 3 | NA |
| 30 | 1 | 2 | NA | NA | NA |
| 34 | 0 | 0 | NA | NA | NA |
| 35 | NA | NA | NA | NA | NA |
| 36 | NA | NA | NA | NA | NA |
| 37 | 2 | 3 | 3 | NA | 3 |
| 38 | 0 | 3 | 1 | 1 | 0 |
| 39 | NA | NA | NA | NA | NA |
| 40 | 1 | 0 | 0 | NA | 0 |
| 41 | NA | NA | NA | NA | NA |
| 45 | NA | NA | NA | NA | NA |
| 48 | 1 | NA | 1 | NA | NA |
| 49 | NA | NA | NA | NA | NA |
| 50 | NA | NA | NA | NA | NA |
| 51 | 2 | NA | NA | NA | 0 |
| 52 | 0 | 1 | 0 | 0 | 0 |
| 53 | 2 | 3 | NA | NA | NA |
| 54 | NA | NA | NA | NA | NA |
| 55 | NA | NA | NA | NA | NA |
| 56 | NA | NA | NA | NA | NA |
| 58 | NA | NA | NA | NA | NA |
| 59 | 0 | NA | NA | 0 | NA |
| 60 | NA | NA | NA | NA | NA |
| 61 | 0 | 0 | 0 | NA | NA |
| 68 | NA | NA | NA | NA | NA |
| 81 | 0 | 0 | NA | NA | NA |
| 82 | NA | NA | NA | NA | NA |
| 84 | NA | NA | NA | NA | NA |
| 86 | NA | NA | NA | NA | NA |
| 101 | NA | NA | NA | NA | NA |
| 102 | NA | NA | NA | NA | NA |

**Supplementary Table 1: AMDP ratings for amotivation across visits**

AMDP clinician ratings for amotivation were obtained at baseline and at follow-up assessments at 3, 6, 9, and 12 months. Ratings used a 0–3 scale (0 = absent, 1 = mild, 2 = moderate, 3 = severe; higher scores indicate greater severity). “NA” denotes time points with no AMDP rating available (e.g., no outpatient visit or no subsequent hospitalization).

| **Subject ID** | **0 months** | **3 months** | **6 months** | **9 months** | **12 months** |
| --- | --- | --- | --- | --- | --- |
| 5 | 1 | 1 | NA | NA | NA |
| 8 | 0 | 0 | NA | 0 | NA |
| 9 | 0 | 0 | NA | 0 | NA |
| 10 | 1 | 0 | NA | NA | NA |
| 11 | 0 | NA | NA | 1 | 0 |
| 12 | 1 | NA | NA | 0 | 2 |
| 13 | NA | NA | NA | NA | NA |
| 14 | NA | NA | NA | NA | NA |
| 15 | NA | NA | NA | NA | NA |
| 16 | NA | NA | NA | NA | NA |
| 17 | 1 | 1 | 1 | NA | 1 |
| 19 | NA | NA | NA | NA | NA |
| 20 | NA | NA | NA | NA | NA |
| 21 | 2 | 2 | 2 | NA | NA |
| 22 | 3 | 3 | NA | NA | 3 |
| 23 | 1 | 0 | NA | 0 | NA |
| 24 | 0 | 0 | 0 | 0 | 0 |
| 25 | NA | NA | NA | NA | NA |
| 26 | NA | NA | NA | NA | NA |
| 27 | NA | NA | NA | NA | NA |
| 28 | 0 | NA | 0 | NA | 0 |
| 29 | 1 | 1 | NA | 2 | NA |
| 30 | 0 | 2 | NA | NA | NA |
| 34 | 0 | 0 | NA | NA | NA |
| 35 | NA | NA | NA | NA | NA |
| 36 | NA | NA | NA | NA | NA |
| 37 | 2 | 2 | 2 | NA | 3 |
| 38 | 0 | 3 | 1 | 2 | 2 |
| 39 | NA | NA | NA | NA | NA |
| 40 | 0 | 0 | 0 | NA | 0 |
| 41 | NA | NA | NA | NA | NA |
| 45 | NA | NA | NA | NA | NA |
| 48 | 1 | NA | 1 | NA | NA |
| 49 | NA | NA | NA | NA | NA |
| 50 | NA | NA | NA | NA | NA |
| 51 | 2 | NA | NA | NA | 1 |
| 52 | 0 | 1 | 0 | 0 | 0 |
| 53 | 0 | 2 | NA | NA | NA |
| 54 | NA | NA | NA | NA | NA |
| 55 | NA | NA | NA | NA | NA |
| 56 | NA | NA | NA | NA | NA |
| 58 | NA | NA | NA | NA | NA |
| 59 | 1 | NA | NA | 0 | NA |
| 60 | NA | NA | NA | NA | NA |
| 61 | 0 | 0 | 0 | NA | NA |
| 68 | NA | NA | NA | NA | NA |
| 81 | 0 | 0 | NA | NA | NA |
| 82 | NA | NA | NA | NA | NA |
| 84 | NA | NA | NA | NA | NA |
| 86 | NA | NA | NA | NA | NA |
| 101 | NA | NA | NA | NA | NA |
| 102 | NA | NA | NA | NA | NA |

**Supplementary Table 2: AMDP ratings for social withdrawal across visits**

Conventions as in Supplementary Table 1.

| **Subject ID** | **0 months** | **3 months** | **6 months** | **9 months** | **12 months** |
| --- | --- | --- | --- | --- | --- |
| 5 | 1 | 0 | NA | NA | NA |
| 8 | 0 | 0 | NA | 0 | NA |
| 9 | 0 | 0 | NA | 0 | NA |
| 10 | 2 | 0 | NA | NA | NA |
| 11 | 0 | NA | NA | 0 | 0 |
| 12 | 0 | NA | NA | 0 | 0 |
| 13 | NA | NA | NA | NA | NA |
| 14 | NA | NA | NA | NA | NA |
| 15 | NA | NA | NA | NA | NA |
| 16 | NA | NA | NA | NA | NA |
| 17 | 1 | 1 | 0 | NA | 1 |
| 19 | NA | NA | NA | NA | NA |
| 20 | NA | NA | NA | NA | NA |
| 21 | 2 | 2 | 1 | NA | NA |
| 22 | 2 | 2 | NA | NA | 2 |
| 23 | 1 | 0 | NA | 0 | NA |
| 24 | 0 | 0 | 0 | 0 | 0 |
| 25 | NA | NA | NA | NA | NA |
| 26 | NA | NA | NA | NA | NA |
| 27 | NA | NA | NA | NA | NA |
| 28 | 0 | NA | 1 | NA | 0 |
| 29 | 1 | 0 | NA | 1 | NA |
| 30 | 1 | 2 | NA | NA | NA |
| 34 | 1 | 0 | NA | NA | NA |
| 35 | NA | NA | NA | NA | NA |
| 36 | NA | NA | NA | NA | NA |
| 37 | 2 | 3 | 3 | NA | 3 |
| 38 | 0 | 0 | 0 | 0 | 0 |
| 39 | NA | NA | NA | NA | NA |
| 40 | 1 | 0 | 0 | NA | 0 |
| 41 | NA | NA | NA | NA | NA |
| 45 | NA | NA | NA | NA | NA |
| 48 | 0 | NA | 0 | NA | NA |
| 49 | NA | NA | NA | NA | NA |
| 50 | NA | NA | NA | NA | NA |
| 51 | 1 | NA | NA | NA | 0 |
| 52 | 0 | 0 | 0 | 0 | 0 |
| 53 | 0 | 1 | NA | NA | NA |
| 54 | NA | NA | NA | NA | NA |
| 55 | NA | NA | NA | NA | NA |
| 56 | NA | NA | NA | NA | NA |
| 58 | NA | NA | NA | NA | NA |
| 59 | 1 | NA | NA | 0 | NA |
| 60 | NA | NA | NA | NA | NA |
| 61 | 0 | 0 | 0 | NA | NA |
| 68 | NA | NA | NA | NA | NA |
| 81 | 0 | 0 | NA | NA | NA |
| 82 | NA | NA | NA | NA | NA |
| 84 | NA | NA | NA | NA | NA |
| 86 | NA | NA | NA | NA | NA |
| 101 | NA | NA | NA | NA | NA |
| 102 | NA | NA | NA | NA | NA |

**Supplementary Table 3: AMDP ratings for affective constriction across visits**

Conventions as in Supplementary Table 1.

| **Subject ID** | **as 0 months** | **as 3 months** | **as 6 months** | **as 9 months** | **as 12 months** |
| --- | --- | --- | --- | --- | --- |
| 5 | 1 | 0 | NA | NA | NA |
| 8 | 0 | 0 | NA | 0 | NA |
| 9 | 0 | 0 | NA | 0 | NA |
| 10 | 2 | 0 | NA | NA | NA |
| 11 | 0 | NA | NA | 0 | 0 |
| 12 | 0 | NA | NA | 0 | 0 |
| 13 | NA | NA | NA | NA | NA |
| 14 | NA | NA | NA | NA | NA |
| 15 | NA | NA | NA | NA | NA |
| 16 | NA | NA | NA | NA | NA |
| 17 | 0 | 0 | 0 | NA | 0 |
| 19 | NA | NA | NA | NA | NA |
| 20 | NA | NA | NA | NA | NA |
| 21 | 2 | 2 | 1 | NA | NA |
| 22 | 2 | 2 | NA | NA | 2 |
| 23 | 0 | 0 | NA | 0 | NA |
| 24 | 0 | 0 | 0 | 0 | 0 |
| 25 | NA | NA | NA | NA | NA |
| 26 | NA | NA | NA | NA | NA |
| 27 | NA | NA | NA | NA | NA |
| 28 | 0 | NA | 2 | NA | 0 |
| 29 | 1 | 0 | NA | 1 | NA |
| 30 | 1 | 2 | NA | NA | NA |
| 34 | 1 | 0 | NA | NA | NA |
| 35 | NA | NA | NA | NA | NA |
| 36 | NA | NA | NA | NA | NA |
| 37 | 1 | 1 | 1 | NA | 1 |
| 38 | 0 | 0 | 0 | 0 | 0 |
| 39 | NA | NA | NA | NA | NA |
| 40 | 1 | 0 | 0 | NA | 0 |
| 41 | NA | NA | NA | NA | NA |
| 45 | NA | NA | NA | NA | NA |
| 48 | 0 | NA | 0 | NA | NA |
| 49 | NA | NA | NA | NA | NA |
| 50 | NA | NA | NA | NA | NA |
| 51 | 1 | NA | NA | NA | 0 |
| 52 | 0 | 0 | 0 | 0 | 0 |
| 53 | 0 | 2 | NA | NA | NA |
| 54 | NA | NA | NA | NA | NA |
| 55 | NA | NA | NA | NA | NA |
| 56 | NA | NA | NA | NA | NA |
| 58 | NA | NA | NA | NA | NA |
| 59 | 0 | NA | NA | 0 | NA |
| 60 | NA | NA | NA | NA | NA |
| 61 | 0 | 0 | 1 | NA | NA |
| 68 | NA | NA | NA | NA | NA |
| 81 | 1 | 1 | NA | NA | NA |
| 82 | NA | NA | NA | NA | NA |
| 84 | NA | NA | NA | NA | NA |
| 86 | NA | NA | NA | NA | NA |
| 101 | NA | NA | NA | NA | NA |
| 102 | NA | NA | NA | NA | NA |

**Supplementary Table 4: AMDP ratings for affective rigidity across visits**

Conventions as in Supplementary Table 1.

***Model Comparison***

We performed model comparison between the 7-, 6-, and 4-parameter models using Leave-One-Out Information Criterion (LOOIC) scores to assess model fit. Lower LOOIC scores indicate better model performance. The scores for each model across four task conditions are reported below.

| **Conditions** | **7 param model**  **(**$\boldsymbol{\alpha}_{\boldsymbol{1}}\boldsymbol{,}\boldsymbol{\alpha}_{\boldsymbol{2}}\boldsymbol{,}\boldsymbol{\beta}_{\boldsymbol{1}}\boldsymbol{,}\boldsymbol{\beta}_{\boldsymbol{2}}\boldsymbol{, p, \lambda, \omega)}$  **(LOOIC)** | **6 param model**  **(**$\boldsymbol{\alpha}_{\boldsymbol{1}}\boldsymbol{,}\boldsymbol{\alpha}_{\boldsymbol{2}}\boldsymbol{,}\boldsymbol{\beta}_{\boldsymbol{1}}\boldsymbol{,}\boldsymbol{\beta}_{\boldsymbol{2}}\boldsymbol{, p, \omega)}$  **(LOOIC)** | **4 param model**  **(**$\boldsymbol{\alpha, \beta, p, \omega)}$  **(LOOIC)** |
| --- | --- | --- | --- |
| HC reward | 21698.03 | 21744.64 | 22437.92 |
| SSD reward | 21983.22 | 22137.92 | 22638.50 |
| HC loss | 21768.15 | 21815.54 | 22547.71 |
| SSD loss | 22656.17 | 22693.06 | 23023.83 |

**Supplementary Table 5:** **Model comparison results using (LOOIC) scores for the 7-, 6-, and 4-parameter models.** We found across all four task conditions in healthy controls and in individuals with schizophrenia spectrum disorder, the seven parameters model to be a better fit to the data, indicated by the lower LOOIC scores.

| **Harvard-Oxford Probabilistic Atlas labels** | x | y | z | Peak Stat | Cluster Size |
| --- | --- | --- | --- | --- | --- |
| 49% Temporal Pole, 3% Middle Temporal Gyrus, anterior division, 3% Inferior Temporal Gyrus, anterior division | -54.5 | 5.5 | -36.5 | -3.935 | 200 |
| 56% Middle Temporal Gyrus, anterior division, 12% Inferior Temporal Gyrus, anterior division, 5% Middle Temporal Gyrus, posterior division, 4% Inferior Temporal Gyrus, posterior division | -54.5 | -4.5 | -30.5 | -3.299 | * |
| 25% Cingulate Gyrus, posterior division, 11% Cingulate Gyrus, anterior division | -4.5 | -16.5 | 35.5 | -3.886 | 136 |
| 60% Temporal Occipital Fusiform Cortex, 23% Lingual Gyrus | 27.5 | -44.5 | -12.5 | 3.480 | 136 |
| 49% Occipital Fusiform Gyrus, 6% Lingual Gyrus, 1% Lateral Occipital Cortex, inferior division | -26.5 | -74.5 | -6.5 | 3.140 | 112 |

**Supplementary Table 6: Voxel clusters showing peak MNI coordinates where individuals with SSD exhibited greater BOLD activation in one context relative to the other during model-based decision making.** Positive stat regions show the regions, where the reward activation was higher, while negative values show regions where the loss activation was higher. Negative/Positive x coordinates indicate activation in the left/right hemisphere of the brain. The activation cluster labels are derived from the Harvard-Oxford Probabilistic Atlas and are corrected for false positive error at 0.005. Regions with * cluster size belong to the same cluster just above.

| **Harvard-Oxford Probabilistic Atlas labels** | x | y | z | Peak Stat | Cluster Size |
| --- | --- | --- | --- | --- | --- |
| 50% Occipital Pole, 2% Supracalcarine Cortex | -0.5 | -98.5 | 13.5 | -3.125 | 80 |
| 13% Postcentral Gyrus, 12% Superior Parietal Lobule, 4% Supramarginal Gyrus, anterior division, 1% Supramarginal Gyrus, posterior division | -28.5 | -40.5 | 45.5 | 5.663 | 2944 |
| 21% Lateral Occipital Cortex, superior division, 18% Superior Parietal Lobule, 4% Angular Gyrus, 2% Supramarginal Gyrus, posterior division | -24.5 | -58.5 | 43.5 | 3.560 | * |
| 34% Middle Frontal Gyrus, 8% Superior Frontal Gyrus, 5% Frontal Pole | -26.5 | 31.5 | 31.5 | 4.846 | 1024 |
| 66% Occipital Pole | 21.5 | -102.5 | -2.5 | 4.058 | 192 |
| 13% Supramarginal Gyrus, posterior division, 12% Angular Gyrus, 1% Lateral Occipital Cortex, superior division | -40.5 | -52.5 | 33.5 | 3.797 | 552 |
| 44% Angular Gyrus, 8% Supramarginal Gyrus, posterior division, 7% Lateral Occipital Cortex, superior division | -48.5 | -56.5 | 27.5 | 3.171 | * |
| 47% Cingulate Gyrus, anterior division, 45% Paracingulate Gyrus, | -4.5 | 29.5 | 27.5 | 3.778 | 392 |
| 37% Right Cerebral Cortex, 11% Planum Temporale, 9% Heschl's Gyrus (includes H1 and H2), 6% Insular Cortex | 33.5 | -28.5 | 11.5 | 3.696 | 160 |
| 42% Superior Parietal Lobule, 9% Lateral Occipital Cortex, superior division | -26.5 | -54.5 | 55.5 | 3.660 | 176 |
| 19% Lateral Occipital Cortex, superior division, 1% Angular Gyrus | 33.5 | -60.5 | 33.5 | 3.502 | 112 |
| 38% Superior Frontal Gyrus, 6% Middle Frontal Gyrus | -20.5 | 1.5 | 57.5 | 3.440 | 264 |
| 39% Superior Temporal Gyrus, posterior division, 36% Middle Temporal Gyrus, posterior division, 5% Middle Temporal Gyrus, temporooccipital part, 4% Supramarginal Gyrus, posterior division | -62.5 | -38.5 | 1.5 | 3.402 | 544 |
| 57% Middle Temporal Gyrus, temporooccipital part, 17% Middle Temporal Gyrus, posterior division, 8% Angular Gyrus, 5% Left Cerebral White Matter, 3% Superior Temporal Gyrus, posterior division, 2% Supramarginal Gyrus, posterior division | -66.5 | -46.5 | -0.5 | 3.273 | * |
| 70% Cingulate Gyrus, anterior division, 22% Paracingulate Gyrus | -4.5 | 39.5 | 13.5 | 3.357 | 264 |
| 11% Frontal Medial Cortex, 6% Paracingulate Gyrus, 1% Frontal Pole, 1% Cingulate Gyrus, anterior division | -14.5 | 45.5 | -6.5 | 3.301 | 80 |
| 42% Precuneus Cortex, 7% Supracalcarine Cortex, 1% Cuneal Cortex | -8.5 | -62.5 | 25.5 | 3.268 | 192 |
| 5% Frontal Pole, 4% Left Cerebral Cortex | -18.5 | 53.5 | 3.5 | 3.258 | 88 |
| 36% Precentral Gyrus, 2% Postcentral Gyrus | -32.5 | -18.5 | 61.5 | 3.235 | 120 |
| 78% Frontal Pole, 4% Inferior Frontal Gyrus, pars triangularis | -44.5 | 41.5 | 5.5 | 3.213 | 136 |
| 49% Middle Frontal Gyrus, 7% Inferior Frontal Gyrus, pars triangularis, 3% Inferior Frontal Gyrus | -48.5 | 23.5 | 29.5 | 3.207 | 120 |
| 43% Supramarginal Gyrus, anterior division, 17% Supramarginal Gyrus, posterior division, 6% Superior Parietal Lobule, 2% Postcentral Gyrus | -52.5 | -38.5 | 49.5 | 3.174 | 96 |
| 90% Frontal Medial Cortex | 1.5 | 47.5 | -18.5 | 3.148 | 112 |
| 69% Right Thalamus | 13.5 | -28.5 | 13.5 | 3.143 | 136 |

**Supplementary Table 7: Voxel clusters showing peak MNI coordinates, where healthy controls exhibited greater BOLD activation in one context relative to the other during model-based decision making.** Positive stat regions show the regions, where the reward activation was higher, while negative values show regions where the loss activation was higher. Negative/Positive x coordinates indicates activation in the left/right hemisphere of the brain. The activation cluster labels are derived from the Harvard-Oxford Probabilistic Atlas and are corrected for false positive error at 0.005. Regions with * cluster size belong to the same cluster just above.
